# Heterogeneity in deep brain stimulation induced gamma oscillations may be explained by bifurcations in neural dynamics

**DOI:** 10.64898/2026.02.12.26346178

**Authors:** Stanisław W. Biber, James J. Sermon, Jonathan Kaplan, Johannes L. Busch, Andrea A. Kühn, Derk-Jan Dijk, Timothy Denison, Anne C. Skeldon

## Abstract

**Objective:** Oscillations in field potentials are associated with a large spectrum of brain function. These oscillations are altered by deep brain stimulation (DBS), but the mechanistic understanding of interactions between brain oscillations and DBS is incomplete. In people living with Parkinson’s disease, DBS can alter pre-existing gamma oscillations or induce gamma oscillations, with a frequency at half the stimulation frequency in both cases. However, these half-harmonic responses, vary between patients. The objective of this work is to provide a dynamical-systems explanation for the heterogeneity in response to DBS.

**Approach:** A Wilson-Cowan model of neuronal populations was used to provide a broad theoretical framework that could explain the heterogeneity of observed responses.

**Main result:** In the absence of stimulation and noise, the model exhibited either damped oscillations or self-sustained oscillations, depending on parameter values. Off-stimulation behaviour determined observed stimulation response. When oscillations in the gamma range were strongly-damped off-stimulation, the only observed stimulation response was a driven oscillation at the stimulation frequency. When oscillations were weakly-damped off-stimulation, stimulation resulted in either harmonic or half-harmonic responses. Half-harmonic responses only occurred for a band of frequencies around half the stimulation frequency and only for sufficiently large amplitude stimulation. When self-sustained oscillations were present off-stimulation, the self-sustained oscillations were entrained by the stimulation frequency, and harmonic, half-harmonic and many other subharmonic responses emerged. Ramping stimulation amplitude up and down revealed hysteresis with the onset and offset of half-harmonic responses appearing at different amplitude thresholds. Addition of noise blurred distinctions between different states but the underlying dynamical structure was preserved.

**Significance:** This framework reproduces the heterogeneity of DBS responses seen in patients and proposes subharmonic phenomena which have not previously been reported. The modelled hysteresis highlights potential challenges for adaptive control systems which rely on a one-to-one relationship between stimulation settings and stimulation outcome.

## 1 Introduction

Deep brain stimulation (DBS) is an invasive neurostimulation technique used in the treatment of neurological conditions where pharmacological treatments prove to be ineffective. Predominantly used for the treatment of Parkinson’s Disease (PD), said to be the fastest growing neurological condition [13], the number of DBS devices implanted each year is increasing [39].

The approach to the delivery of DBS treatment has hardly changed since its inception in the 1980s [24, 23, 19].. The default method continues to rely on delivery of electrical pulses at a stimulation setting (pulse width, waveform, amplitude and frequency) chosen by clinicians and typically fixed at the same value throughout the day and night [51, 25, 29]. The location of the pulse delivery depends on the neurological condition, with subthalamic nucleus (STN) or globus pallidus (GPi) the most common locations used in the treatment of PD [54].

State-of-the-art DBS devices have considerable capabilities to simultaneously deliver stimulation and monitor local field potentials (LFP) at the point of stimulation. This dual capability has led to adaptive DBS systems which adjust stimulation settings in real time in response to live measurements of the LFP [27, 33]. To better exploit the full capabilities of these devices a fuller understanding of the mechanisms behind DBS is needed.

According to a widely accepted concept, the effects of DBS may be mediated by altering oscillations in neural networks [10]. For example, in Parkinson’s, DBS suppresses beta oscillations, which are associated with bradykinesia and rigidity [7, 2]. In addition, DBS can induce or interact with oscillations in the gamma band. Spontaneous gamma oscillations have a frequency between 60-90Hz. Gamma oscillations in response to DBS are characterised by narrowband activity with a width of approximately 5-10 Hz. These are referred to as finely tuned gamma (FTG) oscillations. They occur at half the stimulation frequency i.e. one neuronal oscillation for every two DBS pulses [28, 46, 32], and have been observed in recordings from the points of stimulation (STN or GPi), as well as from electrocorticograph (ECoG) and electroencephalogram (EEG) recordings [41, 22, 52]. These half-harmonic responses are not seen in all patients and were initially thought to be a device-induced artefact but are now understood as a physiological response of the brain [40, 30].

In some patients, even in the absence of stimulation, narrowband gamma activity is observed in response to dopaminergic medication [52, 30]. This dopaminergic gamma-band response is usually referred to as spontaneous finely tuned gamma (sFTG). DBS can shift the frequency of the sFTG oscillation so that the peak frequency of response is at precisely half the stimulation frequency, i.e. a half-harmonic response. Thus, responses in the gamma band are heterogeneous between patients: half-harmonic FTG responses are found in some patients, but not others. In some patients, these half-harmonic responses occur as a result of shifting sFTG oscillations, but not in all cases [30].

The brain’s rhythmic nature has been extensively studied and described [9], with references to it as a neuronal oscillator dating back to the 1980s [44]. Physiologically-based mathematical and computational models, to account for the oscillatory phenomena related to DBS, have been proposed [26], but as yet there is no consensus as to the key populations / networks involved. For example, Rubin & Terman considered the interaction of subthalamic nuclei (STN), the internal and external segments of the globus pallidus (GPi and GPe respectively) and thalamocortical (TC) cells [37]. Their framework was then extended to include multi-site STN stimulation in [17]. More recently Ahmadipour et al. considered interactions between oscillations in the striatum network and the globus pallidus pars externa, and the interaction with the STN [1]. Despite substantial progress there remains a disconnect between current physiological models and practical guidance for engineers designing control systems and clinicians delivering therapy to patient.

An alternative approach is to consider phenomenological models which focus on general principles which are agnostic to the details of the physiology [31, 43]. For example, the alteration of sFTG rhythms by DBS has been described using a Wilson-Cowan modelling framework [41, 14], with one excitatory and one inhibitory population of neurons. In [41], the stimulation pulse was modelled as a forcing term acting on the inhibitory neurons. Model parameters were found by fitting to off-stimulation patient data. Predictions were then made as to the effect of stimulation, successfully demonstrating that stimulation could result in the observed half-harmonic responses. The modelling made the highly non-intuitive prediction, subsequently observed in patients, that increasing stimulation amplitude does not necessarily result in a stronger half-harmonic response but can instead lead to the disappearance of the response. The half-harmonic response was linked to the classic Arnold-tongue entrainment framework in which the frequency and amplitude of the response are dependent on the stimulation frequency and amplitude [2]. In the DBS field, the half-harmonic response highlighted in the phenomenological model of Sermon et al. [41, 14] remains a rare example of a model prediction which subsequently was observed in recordings.

Here, our aim is to investigate whether a single modelling framework can provide a mechanistic account of the occurrence, or otherwise, of the different responses to DBS in the gamma band. We start by illustrating that the modelling framework needs to be nonlinear for half-harmonic responses to be observed in response to simulation. This illustration translates a standard result from linear systems theory to the DBS setting. We then turn to the (nonlinear) Wilson-Cowan mathematical formulation and carry out a dynamical-systems analysis highlighting that the model can exhibit either damped or self-sustained oscillations in the absence of stimulation, depending on parameters. We systematically explore the response of the model to stimulation and find that the Wilson-Cowan model provides a candidate mechanism explaining the observed heterogeneity of responses to stimulation, see Fig. 1. We highlight that multiple subharmonic responses, not only the half-harmonic, should occur and identify the presence of hysteresis. This extends the previous formulation, which referred to this phenomenon in terms of Arnold tongues only. We end with a short discussion of the interpretation and significance of our results.

**Figure 1:**
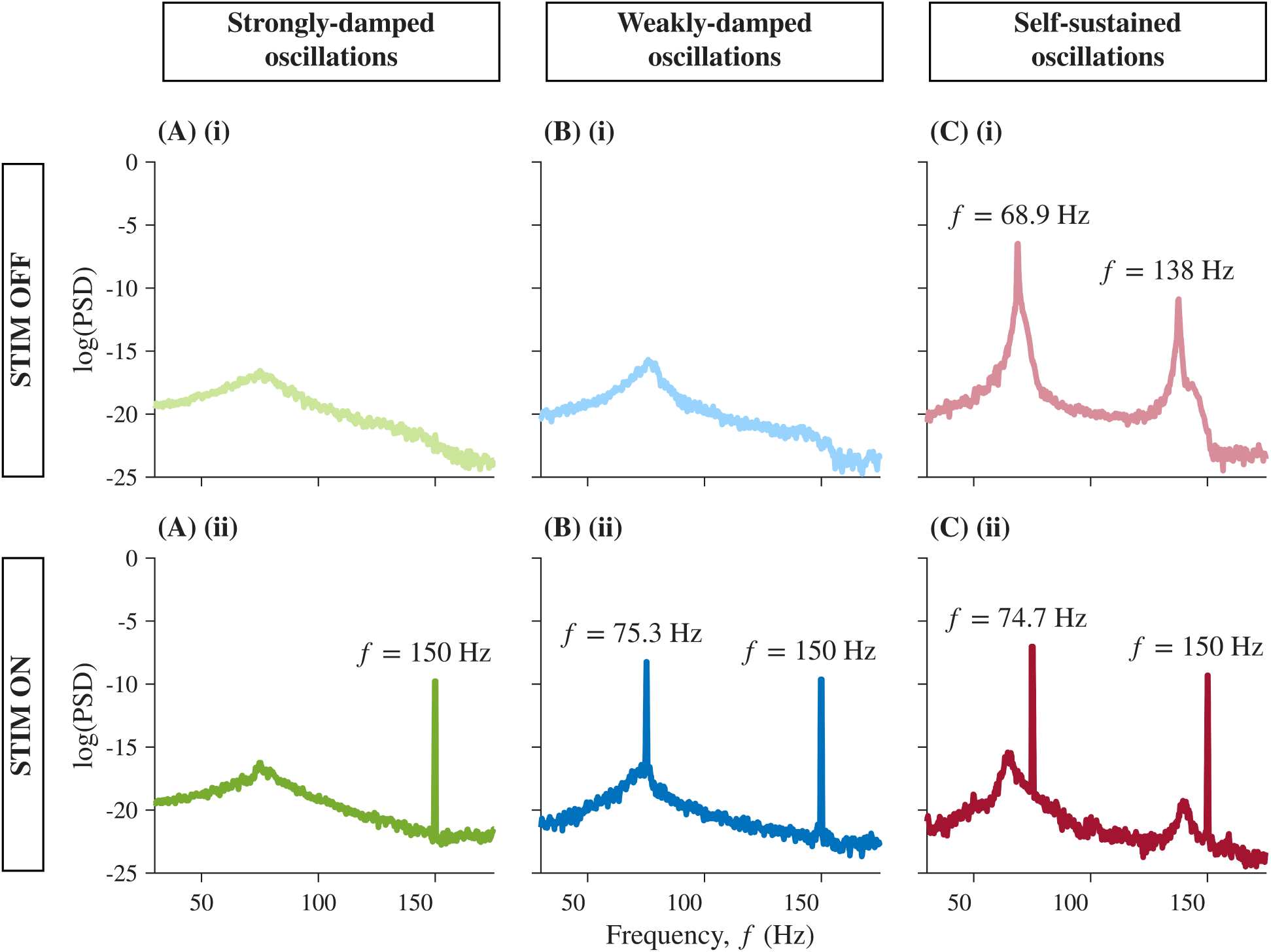
Simulation of the heterogeneity in gamma band responses to deep brain stimulation at 150 Hz. Three simulated scenarios are shown. (A) When endogenous oscillations are strongly damped (no spontaneous finely tuned gamma, sFTG, during stim-off), no half-harmonic responses are present during stim on. (B) When endogenous oscillations are weakly-damped (still no sFTG during stim off), half-harmonic responses can be induced. (C) When endogenous oscillations are self-sustained (i.e. sFTG oscillations are observed) the stimulation shifts the peak frequency to half the stimulation frequency (half-harmonic). All panels show power spectral densities (PSD) of simulated local field potential (LFP) recordings. Top panels: off-stimulation. Bottom panels: stimulation at 150 Hz and an amplitude 10 a.u..

## 2 Methods

### 2.1 Wilson-Cowan Model

We used the Wilson-Cowan model [11, 53], as adapted from [41]. The model is not specific to STN, GPi, or other possible stimulation locations. Instead, it considers the interaction between one excitatory and one inhibitory population of neurons with firing rates at time *t* of *E*(*t*) and *I*(*t*), respectively. The network architecture is shown in Fig. 2(A). Assumptions are: (i) in the absence of input from other neurons, the firing rate of excitatory and inhibitory neurons decay in proportion to their instantaneous firing levels with time constants *τ_E_* and *τ_I_* respectively; (ii) when inhibitory neurons fire sufficiently strongly, they inhibit the firing of excitatory neurons where the strength of the inhibition is determined by the parameter *ω_IE_*; (iii) when excitatory neurons fire sufficiently strongly, they excite the firing of inhibitory neurons where the strength of the excitation is determined by the parameter *ω_EI_*; (iv) excitatory and inhibitory populations are subject to constant background inputs *η_E_* and *η_I_* respectively; (v) excitatory neurons are also self-excitatory, with the strength of the self-excitation determined by *ω_EE_* and (vi) DBS acts by providing a further excitatory input to inhibitory neurons.

**Figure 2:**
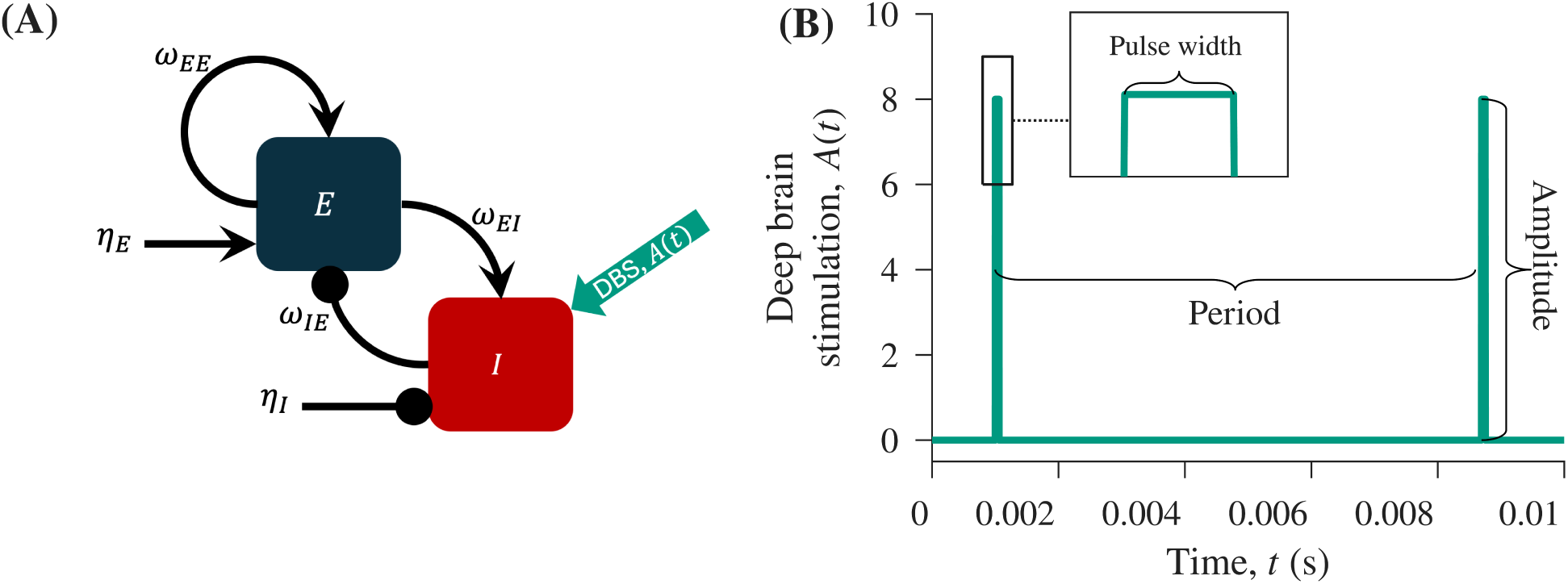
Wilson-Cowan framework for modelling deep brain stimulation. (A) A schematic representation of the Wilson-Cowan model, showing the interaction between excitatory and inhibitory populations with firing rates *E*(*t*) and *I*(*t*), respectively. (B) A graph of the function *A*(*t*), representing the amplitude of the stimulation pulse delivered by the deep brain stimulator as a function of time *t*. Here, the pulse width is set to 6 *×* 10*^−^*^5^s, frequency is 130 Hz, consistent with the standard stimulation settings, and the amplitude is 8 a.u..

Formally, the model is described by

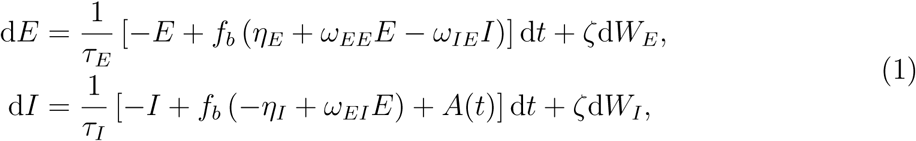

where *f_b_*(*x*) = [1 + exp(*−b*(*x −* 1))]*^−^*^1^ is a logistic function. This logistic function models the fact that inputs to neuronal populations must be sufficiently large to result in large-scale firing of the target neuronal population. For low values of the argument *x*, *f_b_*(*x*) asymptotes to 0, and for high values *f_b_*(*x*) asymptotes to 1. The steepness of the change with *x* is regulated by the parameter *b*, with larger values of *b* corresponding to a steeper change. *A*(*t*) is the DBS forcing term. Stochastic effects are represented by Wiener-processes d*W*_(_*E, I*), with standard deviation *ζ* which is assumed to be the same for both neuronal populations.

We consider the dynamical structure of the equations first in the off-stimulation setting, *A*(*t*) = 0 for all time *t*, and without noise (*ζ* = 0). We then consider the effect of stimulation, *A*(*t*) ≠ 0. The DBS forcing term is taken as a pulse, characterised by its amplitude, frequency, and width, see Fig. 2(B). Results are presented for varying amplitude and frequency. The amplitude is given in terms of arbitrary units as the relation between changes in the firing rates of the excitatory and inhibitory populations and the stimulation amplitude is unknown. Motivated by standard stimulation settings, most results are presented for a pulse width fixed at a constant value (6 *×* 10*^−^*^5^s).

Signals recorded from the brain are inherently ‘noisy’. We therefore also evaluate the extent to which dynamical features on-stimulation are preserved in the presence of a realistic level of noise (*ζ >* 0). The value of *ζ* is set at 0.087, as estimated from patient data in [41].

Generally, for the Wilson-Cowan model, physiologically relevant solutions are considered to have 0 *< E <* 1, 0 *< I <* 1, although noise or a sufficiently large stimulation pulse could in principle knock a solution outside this region. In practice, for the parameter values we used, the stable attractors were sufficiently far from the boundary that solutions remained within the physiological region of relevance. We also note that the unit square in phase space defined by 0 *< E <* 1, 0 *< I <* 1 is globally attracting.

### 2.2 Mathematical methods for studying the behaviour of the Wilson-Cowan model in the absence of stimulation

In the absence of stimulation (and noise), the dynamical behaviour of equations (1) was considered by carrying out a bifurcation analysis. In this approach, the number and stability of steady-state (equilibrium) solutions of the equations are considered, where steady-states correspond to solutions such that the firing rates of excitatory and inhibitory neuronal populations do not change with time (d*E/*d*t* = 0 = d*I/*d*t*). Generically, steady-state solutions are either stable or unstable. Stable steady-state solutions are locally attracting, meaning that when solutions are slightly perturbed, they return to the stable steady-state solution. Unstable steady-state solutions are locally repelling, meaning any small perturbation results in solutions diverging from the unstable steady-state.

We tracked the position of the steady-state solutions as a function of the parameters and considered how their number and stability changed. We identified transition points (bifurcations) where solutions either changed stability or solutions were gained or lost. Of particular interest were points where steady-state solutions lost stability by undergoing a Hopf bifurcation and a periodic solution was created. These periodic solutions (i.e. self-sustained oscillatory solutions) arising at the Hopf bifurcation were also tracked as a function of the parameters.

Understanding the bifurcation structure facilitated an understanding of the overall dynamical behaviour of the model and robustness of the modelling framework. We note that it is possible to find analytical conditions for the steady-state solutions and analytical conditions defining when Hopf bifurcations occur, see the Supplementary Materials Section 1. However, since these conditions are implicit coupled nonlinear equations that are not readily solvable by hand, we turned to numerical bifurcation methods and used the Matcont package for Matlab [12]. Matcont uses numerical continuation methods to find, classify and follow equilibria and periodic solutions of a dynamical system as a function of the parameters and also locates and tracks bifurcation points.

### 2.3 Simulation techniques

In addition to carrying out a numerical bifurcation analysis of the system in the absence of stimulation, we carried out simulations of equations (1), to systematically investigate model behaviour with stimulation-on, simulate DBS neuromodulation experiments, and predict the effect of new stimulation regimes. For deterministic simulations (*ζ* = 0) calculations were carried out by integrating equations (1) using Matlab’s built-in ODE-45 solver. For stochastic simulations (*ζ >* 0), we used the Euler–Maruyama method, with a time step of 10*^−^*^5^ s. All numerical integrations were carried out in Matlab [47, 48].

Choices for the seven physiological parameters (*τ_E_, τ_I_, η_E_, η_I_, ω_EE_, ω_EI_, ω_IE_*) were motivated by the results in [41], in which the Wilson-Cowan model was fitted to patient data, and also by our numerical bifurcation analysis. Three different general physiological parameter regimes were considered corresponding to the existence of (i) strongly-damped oscillations, (ii) weakly-damped oscillations, and (iii) self-sustained oscillations. All baseline physiological parameter values are listed in the Supplementary Materials Section 3.

To systematically investigate model behaviour, equations (1) were integrated for a range of stimulation amplitudes and frequencies and for different initial conditions *E*_0_*, I*_0_. For each setting, the response to the first 500 stimulation pulses was discarded to remove transient behaviour. Following that, the system was integrated for a further 250 stimulation pulses. For each solution the rotation number *ρ* was calculated to classify the type of harmonic or subharmonic response. Here,

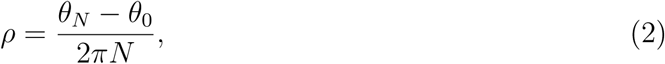

where *θ_N_* is the phase of the oscillator after *N* stimulations and *θ*_0_ is its initial phase. To calculate the oscillator phase *θ_N_* the solutions were first centred at the temporal means of the excitatory ((*E*°) and inhibitory (*I*°) populations (*E*(*t*) *’→ E*(*t*) *− E*°*, I*(*t*) *’→ I*(*t*) *− I*°. The translated solutions in the (*E, I*) plane were then represented using polar coordinates (*r, θ*). The unwrapped value of *θ* after *N* stimulations was used as an estimate of the phase *θ_N_*. When the rotation number was rational (i.e., it could be written as *ρ* = *p/q*, where *p* and *q* are integers), every *q* periods of the forcing results in *p* oscillations of the system. Thus, the first harmonic has a rotation number *ρ* = 1, whereas a half-harmonic response has *ρ* = 1*/*2.

To illustrate the implications of the theoretical framework for experiments, a DBS neuromodulation experiment in which a patient is exposed to DBS at a set frequency. Two different stimulation frequencies were considered, 125 and 150 Hz. The amplitude of the stimulation was ramped up or down in discrete steps, was simulated. As in typical experiments, stimulation amplitude was increased or decreased every 10 seconds, and we chose to change it in steps of 2 a.u.. Numerical solutions of the firing rate of the excitatory population *E*(*t*) were sampled at four-times the stimulation frequency (500 and 600 Hz, respectively). Spectrograms and power spectral densities (PSDs) of the resulting time series were computed using Hamming windows (of 1 s) with no overlap or PSD rescaling.

In some regions of the stimulation frequency-amplitude space, hysteresis was observed. A characteristic of hysteresis is the co-existence of multiple stable solutions to the system. Which solution is observed depends on the initial conditions, with each stable solution having a ‘basin of attraction’. Basins of attraction were constructed by integrating equations (1) over a domain of initial conditions. For each set of initial conditions, the rotation number was computed after 1000 stimulation pulses.

### 2.4 Sensitivity analysis

Robustness of our findings in the case of the deterministic system was examined by performing a sensitivity analysis in which we varied the coupling strength parameters *ω_EE,EI,IE_*, background input parameters *η_E,I_* and the time scale constants *τ_E,I_*. Parameters were varied one at a time by *±*5% from their baseline values.

For each parameter set, we located the relevant equilibrium and calculated the eigenvalues of the local Jacobian. The eigenvalues enabled us to evaluate which side of the Hopf stability boundary the parameters were and, in the damped case, the strength of the damping.

For each parameter set, the model was simulated for varying frequency and amplitude of stimulation as described in section 2.3. In the resulting frequency-amplitude bifurcation set we visually assessed the presence or absence of subharmonic response regions. We then quantitatively focused on the assessment of the half-harmonic tongue. For each frequency-amplitude point, solutions were classified as belonging to the half-harmonic point if the rotation number satisfied *|ρ −* 0.5*| <* 0.02. Regions of complex dynamics, as discussed later, were excluded from the comparison.

For each identified half-harmonic tongue, we estimated the area and the location of the tongue tip in frequency and amplitude dimension. For the case of self-sustained oscillations, we checked for the existence of hysteresis by integrating the system for two initial conditions. Area calculations were expressed relative to the corresponding baseline case. Results were plotted against the perturbation magnitude, with −5%,0%, and 5% representing the decrease, baseline and increase of the parameters, respectively.

## 3 Results

### 3.1 Nonlinearity as a prerequisite for a half-harmonic response to stimulation

Before turning to the Wilson-Cowan framework, we considered if a half-harmonic response could be observed in a dissipative linear system subject to an impulsive periodic stimulation. We assume the system starts from a set initial condition and consider two stimulation pulses at equal time intervals *T*. After two periods (*t* = 2*T*) we assume the system returns to its original position (rotation number *ρ* = 1*/*2) and show that this is only possible if it previously returned to that position at time *t* = *T* (i.e. a harmonic response).

We first sketch a proof for a stable node (one-dimensional, non-oscillatory) case.

Consider a one-dimensional linear system with a stable equilibrium. Without loss of generality, we can assume that the equilibrium is at the origin and therefore that

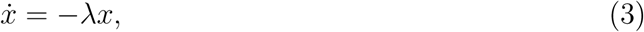

where *λ >* 0. This has a solution of the form *x*(*t*) = *C*e*^−λt^*.

Suppose now that at time *t* = 0 the system is at *x*(0) = *ε* and we apply an instantaneous pulse of size Δ, so that, after the pulse is applied, *x*(0) = *ε* + Δ, and hence *x*(*t*) = (*ε* + Δ)e*^−λt^.* At time *t* = *T* the pulse is applied again, so that after the stimulation *x*(*T*) = (*ε* + Δ)e*^−λT^*+ Δ. After the second pulse, the general form of the solution is *x*(*t*) = [*ε* + Δ + Δe*^λT^*] e*^−λt^*. In particular, if the solution is periodic with period 2*T*, the system returns to the initial position after the two stimulation periods, hence

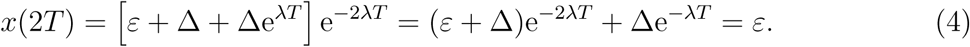

Solving for e*^−λT^*, we see that the only possible solution is 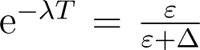. In particular, this means that after one period following the first stimulation pulse (and before the second pulse is applied)

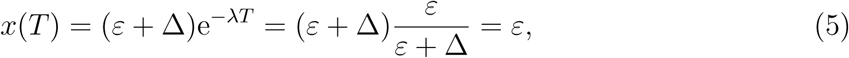

meaning that the system must also return to its original position at *t* = *T*. Hence, the solution is harmonic.

A generalisation of this sketch proof for two-dimensional linear systems is given in Supplementary Materials Section 4. Together, these results imply that a linear system with an impulsive forcing cannot exhibit half-harmonic response in the absence of self-sustained oscillations, and we therefore turn to the (nonlinear) Wilson-Cowan framework.

### 3.2 The Wilson-Cowan framework can exhibit either damped or self-sustained oscillations off-stimulation

In the absence of stimulation (*A*(*t*) = 0), solutions to equations (1) exhibited damped oscillations asymptoting to a stable steady-state value in the firing rates of the neuronal populations for some parameter values, see Fig. 3(A). For other parameter values, the firing rates of the neuronal populations exhibited self-sustained oscillations, see Fig. 3(B).

**Figure 3:**
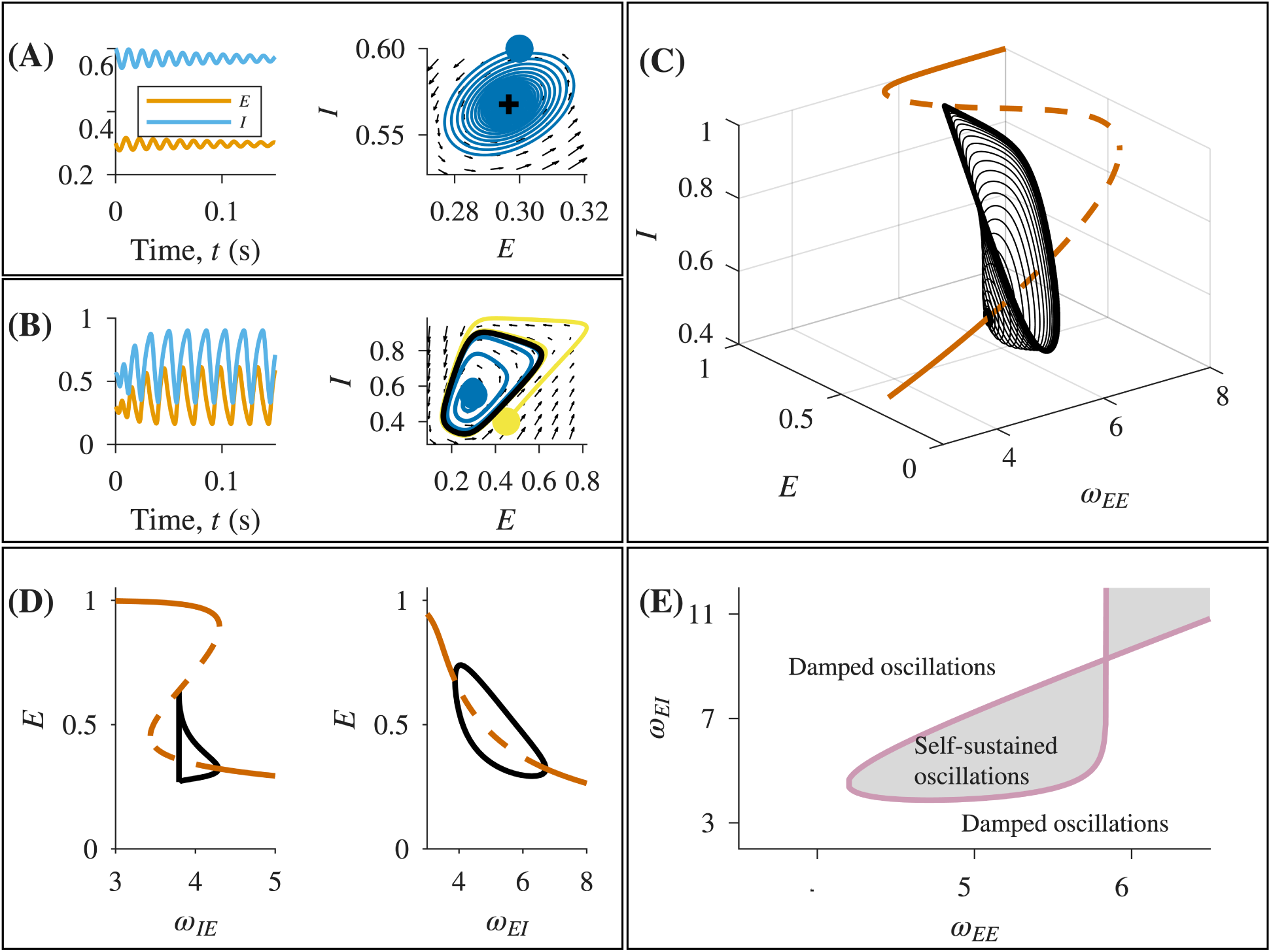
Dynamics and bifurcation structure of the Wilson-Cowan model. Depending on the parameter values, damped oscillations or self-sustained oscillations occurred. (A) Example damped oscillations, here decaying to the stable steady-state *E*(*t*) = 0.30*, I*(*t*) = 0.57 (*ω_EE_* = 4.795). (B) Example self-sustained oscillations (*ω_EE_* = 5.2). In both (A) and (B) the left panel shows the time course of the firing rates of the excitatory (blue) and inhibitory (orange) populations and the right-hand panel shows the corresponding phase plane. On the phase planes, the direction of the flow (small black arrows) and example trajectories (blue and yellow spirals with the initial position marked with a blue or yellow ‘*•*’ sign) are shown. The stable steady-state solution (black ‘+’ sign, panel (A)) and stable self-sustained oscillations (black closed curve, panel (B)) are also shown. (C,D) Bifurcation diagrams showing how steady-state solutions and self-sustained oscillations change as a function of *ω_EE_*(panel C), *ω_IE_*(D, left panel) and *ω_EI_*(D, right panel). In (C) and (D), stable (unstable) steady-states are shown by the brown solid (dashed) curve. In (C) black closed loops represent stable self-sustained oscillatory solutions. In (D), for simplicity, only the firing rate of the excitatory population *E* is shown and the black curves denote the boundary of the periodic orbits. The transition from damped oscillations which decay to a stable steady-state to self-sustained oscillations occurs at Hopf bifurcation points. (E) Bifurcation set showing the position of the Hopf bifurcation points in the *ω_EE_ − ω_EI_* space separating regions of damped oscillations from regions of self-sustained oscillations. Baseline model parameters used for all panels are given in the Supplementary Materials.

To better understand when self-sustained oscillatory behaviour occurs, we considered the behaviour of the system as a function of the model parameters. For example, model behaviour as a function of the strength of the self-excitation of the excitatory population, *ω_EE_*, is shown in Fig. 3(C). For low values of *ω_EE_*, damped oscillations, which decayed to a stable steady-state solution, occurred. On increasing *ω_EE_*, damped oscillations were replaced by self-sustained oscillatory solutions at a Hopf bifurcation. On further increase of *ω_EE_*, self-sustained oscillations were removed by a homoclinic bifurcation in which the oscillatory solution collided with an unstable steady-state solution. For high values of *ω_EE_*, a further stable steady-state solution exists near *E* = 1*, I* = 1 corresponding to high firing rates of both excitatory and inhibitory neurons. We note that for 5.47 *< ω_EE_ <* 7.91 multiple steady-state solutions exist, only one of which is stable.

Similarly, varying other model parameters showed that for some values there were damped oscillations and for other values there were self-sustained oscillations, with the transition between the two states occurring at Hopf bifurcations. For example, see Fig. 3(D, left panel) for the effect of varying the strength of the inhibition of inhibitory neurons on the excitatory neurons *ω_EI_* and Fig. 3(D, right panel) for the effect of varying the strength of the excitation of excitatory neurons on the inhibitory neurons, *ω_IE_*.

Tracking the position of the Hopf bifurcations as a function of the parameters showed how the regions of damped oscillations versus self-sustained oscillations varied in the two parameter *ω_EE_ − ω_EI_* plane, see Fig. 3(E).

### 3.3 The effects of stimulation differed depending on whether off-stimulation oscillations were strongly-damped, weakly-damped or self-sustained

We simulated the Wilson-Cowan model as a function of excitation frequency and excitation amplitude for three different physiological parameter settings corresponding to a regime with strongly-damped oscillations (*ω_EE_* = 4.0), a regime with weakly-damped oscillations (*ω_EE_* = 4.795) and a regime with self-sustained oscillations (*ω_EE_* = 5.2).

#### 3.3.1 The strongly-damped oscillatory parameter regime only exhibited harmonic responses to the driving frequency

Systematic variation of stimulation amplitude and frequency for the strongly-damped case (*ω_EE_* = 4.0) only found harmonic responses for all considered stimulation amplitudes and frequencies, see Supplementary Fig. S3.

#### 3.3.2 The weakly-damped oscillatory parameter regime exhibited both harmonic and half-harmonic responses to the driving frequency

For *ω_EE_*in the weakly-damped regime (*ω_EE_*= 4.795), see Fig. 4 the majority of stimulation settings gave rise to a harmonic response (rotation number *ρ* = 1), corresponding to oscillations driven at the stimulation frequency. For stimulation frequencies around 150-160 Hz and for sufficiently large amplitudes, a half-harmonic response (*ρ* = 1*/*2) was found. No other subharmonic responses were found.

**Figure 4:**
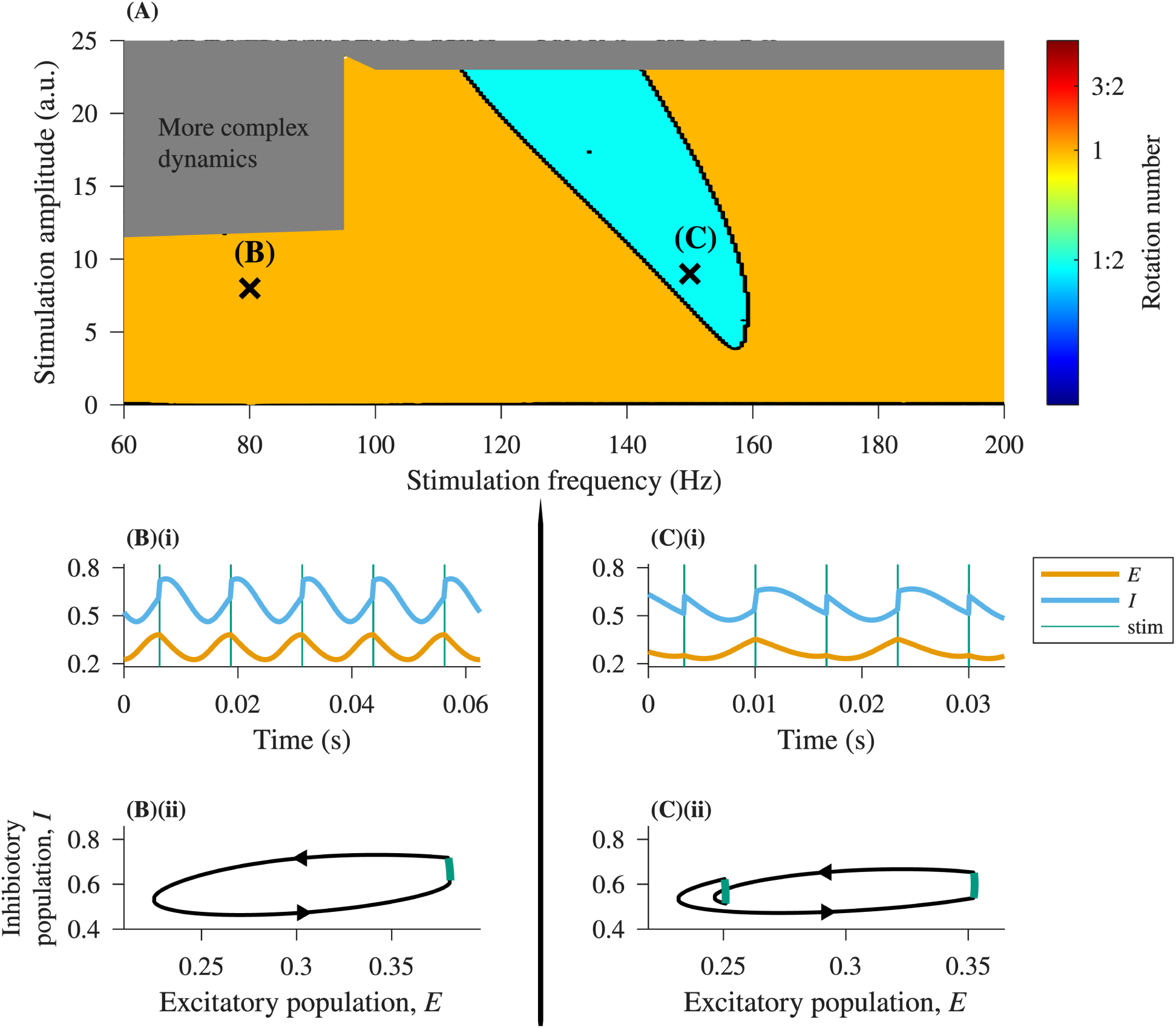
Response to stimulation for a parameter regime with weakly-damped oscillations as a function of stimulation frequency and amplitude. (A) Varying stimulation parameters resulted in either harmonic (orange) or half-harmonic (cyan) responses. Rotation numbers were calculated from the numerical solution using equation (2). Time series and phase portraits for typical harmonic (B) and half-harmonic responses (C) are also shown. In (B) and (C), (i) show time series of the firing rate of excitatory neurons *E*(*t*) and inhibitory neurons *I*(*t*) with vertical lines indicating the stimulation times and (ii) show the associated phase portraits

#### 3.3.3 The self-sustained oscillation regime exhibited responses which were harmonic or subharmonic to the driving frequency

For *ω_EE_*in a region with self-sustained oscillations (*ω_EE_* = 5.2) systematic variation of stimulation amplitude and stimulation frequency showed that both harmonic and many different subharmonics could occur, see Fig. 5, characteristic of an Arnold tongue structure [3]. The areas of harmonic and half-harmonic solutions were the largest and most prominent. These types of solution resembled those in the weakly-damped case, c.f. Fig. 4(B-C) and Fig. 5(B-C). However, there were also regions corresponding to other rational rotation numbers. For example, Fig. 5(D-E) show solutions with *ρ* = 2*/*3 (three stimulation pulses for every two neuronal cycles) and *ρ* = 3*/*4 (four stimulation pulses for every three neuronal cycles). In addition, a region in which there was coexistence of harmonic and subharmonic solutions was found, which we will return to later.

**Figure 5:**
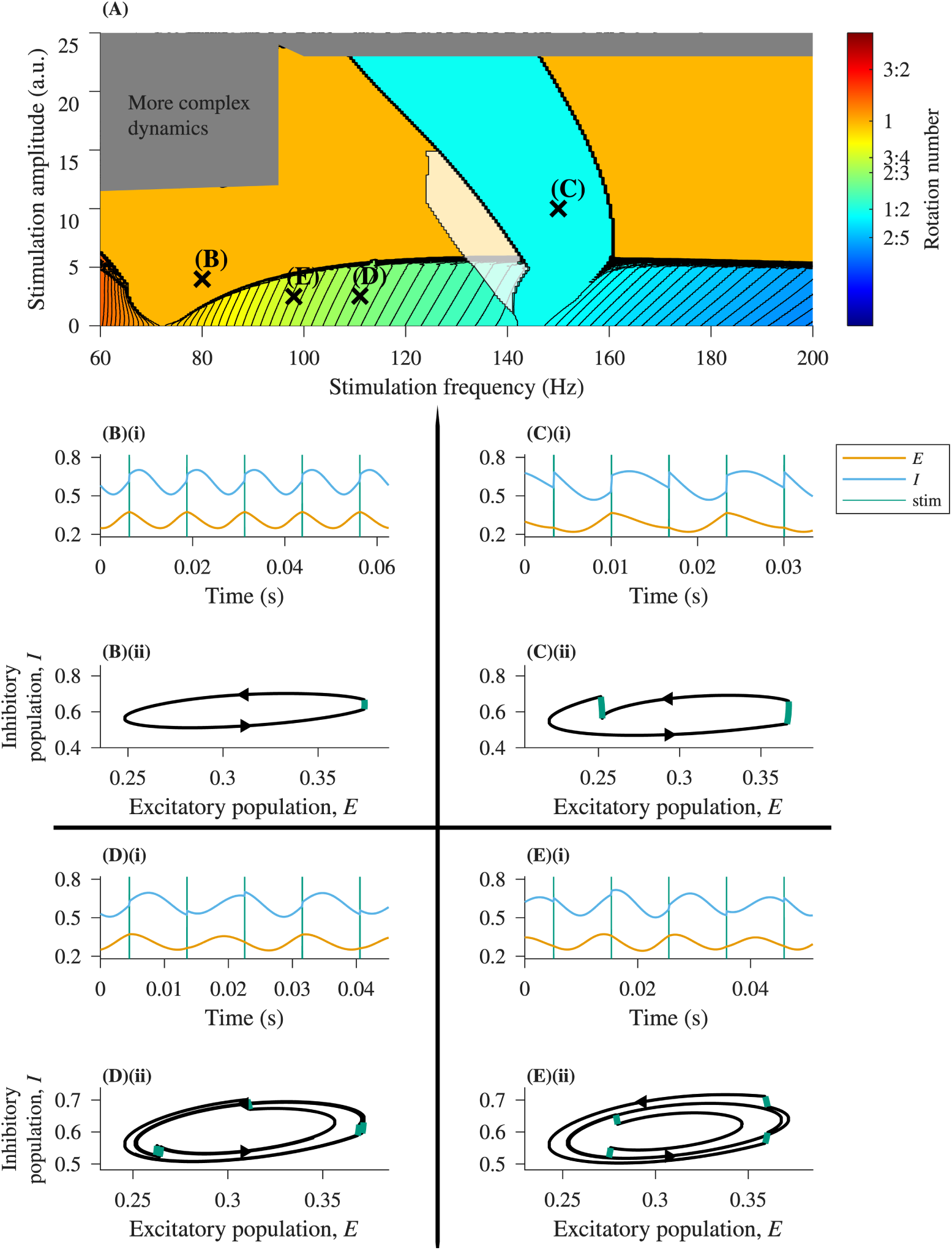
Response to stimulation for a parameter regime with self-sustained oscillations as a function of stimulation frequency and amplitude. (A) Varying stimulation parameters resulted in harmonic response (*ρ* = 1, orange), half-harmonic (*ρ* = 1*/*2, cyan) and many subharmonics (shaded according to rotation number, see the colour bar). Rotation numbers were calculated from the numerical solution using equation (2). Contours were drawn at fixed values of the rotation number, with finer spacing below *ρ* = 1*/*2 to resolve the structure around the half-harmonic response. The white transparent region is a region of coexistence of harmonic and subharmonic responses. Time series and phase portraits for typical responses are shown in (B)-(E). (B) Harmonic response (*ρ* = 1). (C) Half-harmonic response (*ρ* = 1*/*2). (D) Subharmonic response with *ρ* = 2*/*3. (E) Subharmonic response with *ρ* = 3*/*4. In each case, (i) shows the time series of the firing rate of excitatory neurons *E*(*t*) and inhibitory neurons *I*(*t*) and (ii) associated phase portraits.

At large amplitudes, characteristic of Arnold tongue systems [5], more complex dynamics were found. For example, period doubling in which the harmonic orbit doubled its period so that the system oscillated twice for each two stimulation periods, but its orbits are not exactly aligned (this is sometimes referred to as a 2:2 solution). Numerical evaluation of 2:2 solutions still appear as a rotation number of *ρ* = 1.

The results shown in Fig. 5 are for a pulse width fixed at a constant value (6 *×* 10*^−^*^5^ s, the standard stimulation setting). The position and shape of the regions of entrainment depend on the width and shape of the pulse, see Supplementary Materials Figure S4. In all cases multiple subharmonics occurred, not only the half-harmonic.

### 3.4 Simulated DBS neuromodulation experiments suggest observed responses will differ depending on whether off-stimulation oscillations are damped or self-sustained

Simulations of DBS neuromodulation experiments in which DBS stimulation amplitude is ramped up in discrete steps showed different results depending on whether off-stimulation oscillations were damped or self-sustained.

In the regime with strongly-damped oscillations off-stimulation, at all values of the stimulation amplitude only harmonic responses at the stimulation frequency were found, see Fig. 6 (A,B). These observations are consistent with taking a vertical slice up through the stimulation frequency-amplitude plane shown in Supplementary Materials, Fig. S3.

**Figure 6:**
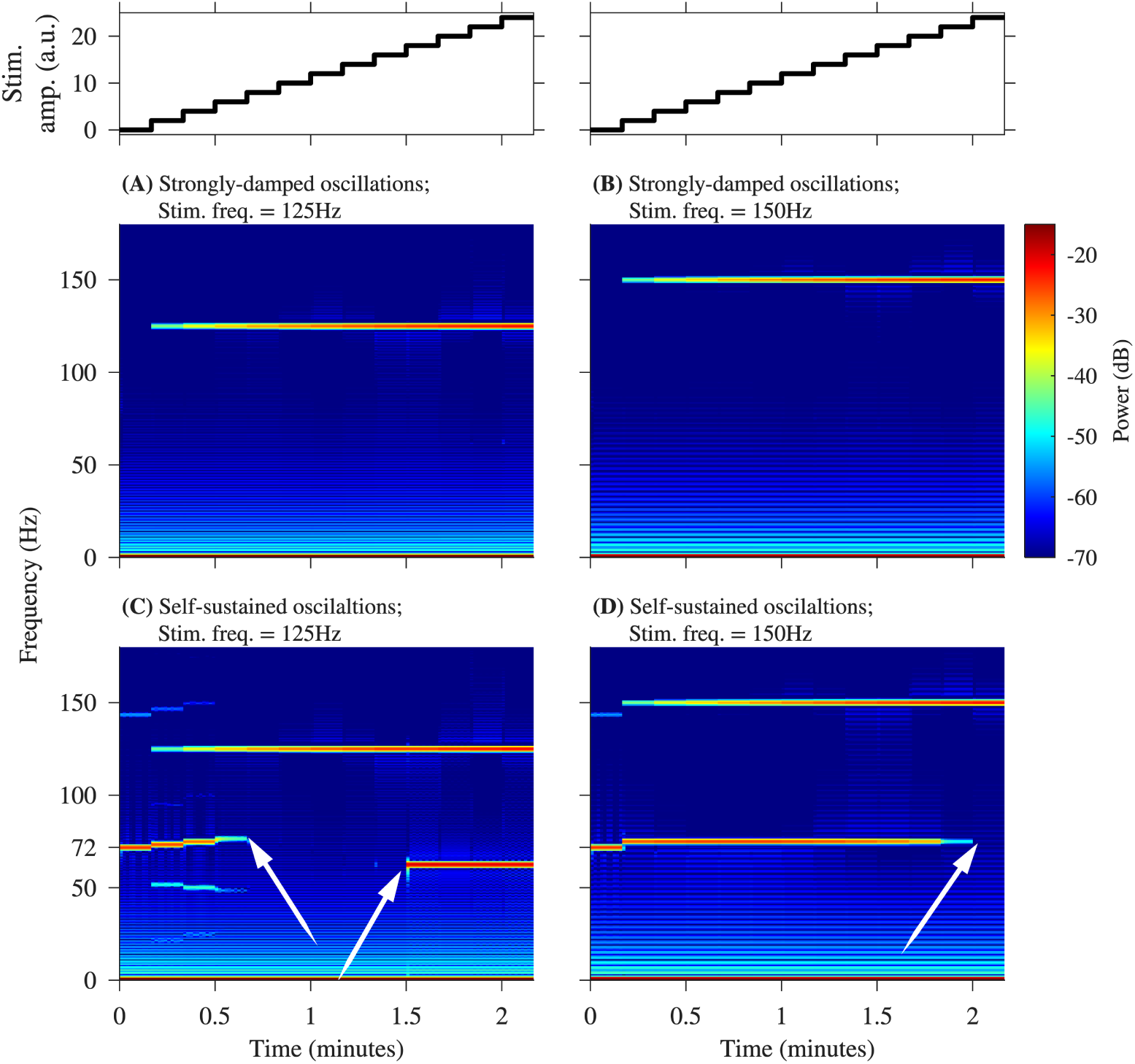
Example predictions of a DBS neuromodulation experiment. Predicted results of stimulation depend on whether oscillations were damped or self-sustained before stimulation was applied. Top panels show the time course of the stimulation amplitude as it is ramped up in discrete steps every 10 seconds (identical panels on the left and right). (A,B) For strongly-damped oscillations (*ω_EE_* = 4.0), only responses at the stimulation frequency were seen. (C,D) In the presence of self-sustained oscillations (*ω_EE_* = 5.2), here with a natural frequency of 72 Hz, various subharmonics were observed depending on stimulation frequency. Arrows indicate points where there is transition between harmonic and subharmonic responses. (A)-(D) all show spectrograms. Left panels (A,C): stimulation frequency 125 Hz. Right panels (B, D): stimulation frequency 150 Hz. Baseline physiological parameter values are given in the Supplementary Materials.

In the regime with weakly-damped oscillations off-stimulation, harmonic responses occurred for both low amplitudes and for high amplitudes of stimulation. But for an intermediate range of stimulation amplitudes, a half-harmonic response was seen, see Supplementary Fig. S5. The stimulation amplitudes for which half-harmonic responses occurred depended on stimulation frequency (cf. left and right panels of Fig. S5). These observations are consistent with the parameter plane shown in Fig. 4 (A), in which half-harmonic responses were dependent on stimulation frequency, only occurred for sufficiently high stimulation amplitude and were lost at large stimulation amplitudes.

In the regime with self-sustained oscillations off-stimulation, half-harmonic and multiple different subharmonics occurred. For example, at a stimulation frequency of 125 Hz and low stimulation amplitudes (0.17 *−* 0.67 s), subharmonic responses in the range of 70-80 Hz (rotation number *ρ ≈* 3*/*5) were found. On increasing amplitude, the subharmonic response was lost and replaced first by the harmonic response and then by a half-harmonic response (*ρ* = 1*/*2), see Fig. 6(C). When the stimulation frequency was increased to 150 Hz, the half-harmonic response was observed for low stimulation amplitudes and persisted until large stimulation amplitudes were reached, Fig. 6(D). These results correspond to taking a vertical slice up through Fig. 5(A) at 125 Hz and 150 Hz respectively.

### 3.5 The Wilson-Cowan framework predicts hysteresis in the transition between different oscillatory states when stimulation is ramped up and down

As shown in Fig. 5, we identified a region in the numerical evaluation of the model, in which there was co-existence of a stable harmonic solution and a stable subharmonic solution for the same stimulation parameter settings, see Fig. 7(A) for an example. Therefore, in any given simulation, which solution was observed depended on the initial conditions (*E*(0) = *E*_0_ and *I*(0) = *I*_0_). Consequently, there were some regions in the initial condition *E*_0_ *−I*_0_ space which resulted in harmonic solutions and some regions which resulted in subharmonic solutions, as illustrated in Fig. 7(B). These ‘basins of attraction’ for the different solutions varied depending on the stimulation settings, including pulse width, amplitude, frequency and the pulse offset time, that is, the time after the stimulation starts at which the first pulse was applied.

**Figure 7:**
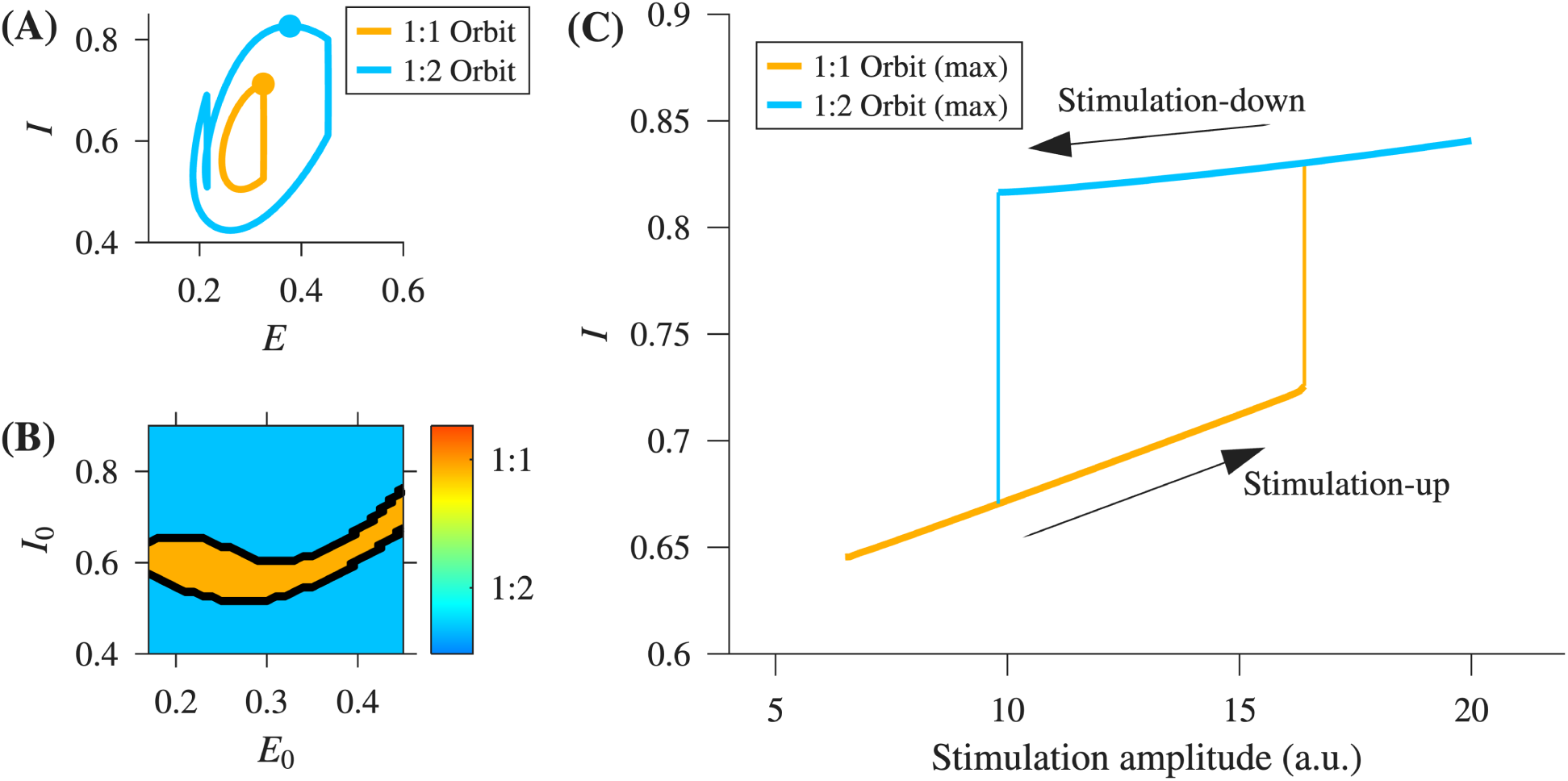
Coexistence of harmonic and subharmonic solutions and resulting hysteresis in transitions on changing stimulation amplitude. (A) Example of co-existing harmonic (orange) and half-harmonic (cyan) solutions. Stimulation amplitude 15 a.u.. Cyan and orange dots ‘*•*’ denote the maximum of the inhibitory population for each orbit. (B) Basins of attraction in the initial condition space for the solutions shown in (A). The orange (cyan) region denotes the set of initial conditions which result in convergence to the harmonic response *ρ* = 1 (half-harmonic response, *ρ* = 1*/*2). (C) Hysteretic loop in harmonic – half-harmonic transitions on ramping stimulation amplitude up and then down. Orange and cyan curves denote the location of the maximum of the harmonic and half-harmonic orbits, respectively. Here, *ω_EE_* = 5.2, stimulation frequency is 125 Hz with an offset of 1/2 of the stimulation period. Other parameters took their baseline values as listed in the Supplementary Materials.

In the model, the coexistence (bistability) of harmonic and subharmonic solutions led to hysteresis in transitions between harmonic and subharmonic solutions on ramping up and ramping down stimulation, see Fig. 7(C). Between stimulation amplitudes of 9.8 and 16.4 a.u. two stable solutions exist. On increasing the simulated stimulation amplitude, the solutions converged to the harmonic solution (*ρ* = 1) until a stimulation amplitude of 16.4 a.u. at which point there was a jump to the half-harmonic solution (*ρ* = 1*/*2). As the simulated stimulation amplitude was decreased, the solutions continue to oscillate with *ρ* = 1*/*2 until the stimulation amplitude reaches 9.8 a.u.

To further illustrate how hysteresis may manifest in DBS neuromodulation experiments in which stimulation amplitude is ramped up and then down, see Fig. 8 for a model prediction. In this numerical example, when the amplitude was increased, a half-harmonic response appeared at a stimulation amplitude of 18 a.u., whereas when the amplitude was decreased, the half-harmonic response persisted until a stimulation amplitude of 10 a.u.. Hysteretic behaviour was found only when self-sustained oscillations were present off-stimulation.

**Figure 8:**
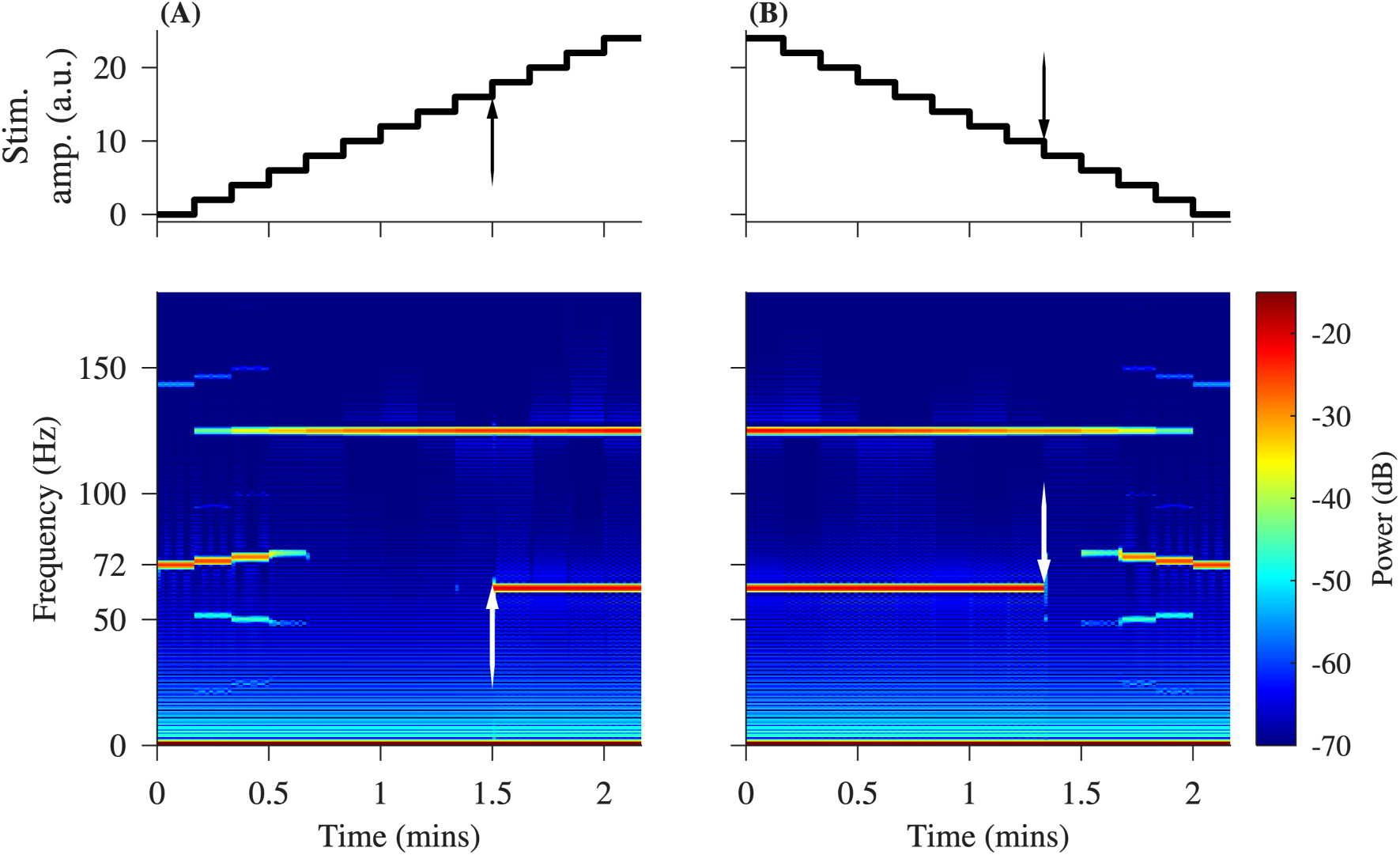
Hysteresis in transitions between different oscillatory states. (A, B) Simulated spectrograms for increasing and decreasing stimulation amplitude respectively. The arrows indicate where the half-harmonic response is gained (A) and (B) lost. Please note the presence of self-sustained oscillations at a frequency of approximately 72 Hz at the beginning (A) and end (B) of the stimulation protocol when the amplitude of stimulation is 0. Here *ω_EE_* = 5.2 and the stimulation frequency was 125 Hz as in Fig. 6(C). All other parameters were set at their baseline parameters as given in the Supplementary Materials.

### 3.6 The effect of background noise on predicted behaviour

To investigate the extent to which observed features were robust to the addition of noise, we repeated simulations with ζ ≠ 0.

Adding noise in general generated broadband weak fluctuations centred on the stable solutions of the noise-free system. These noise-driven fluctuations smeared out sharp features observed in the absence of noise, see Fig. 9 for example PSD plots with and without background noise.

**Figure 9:**
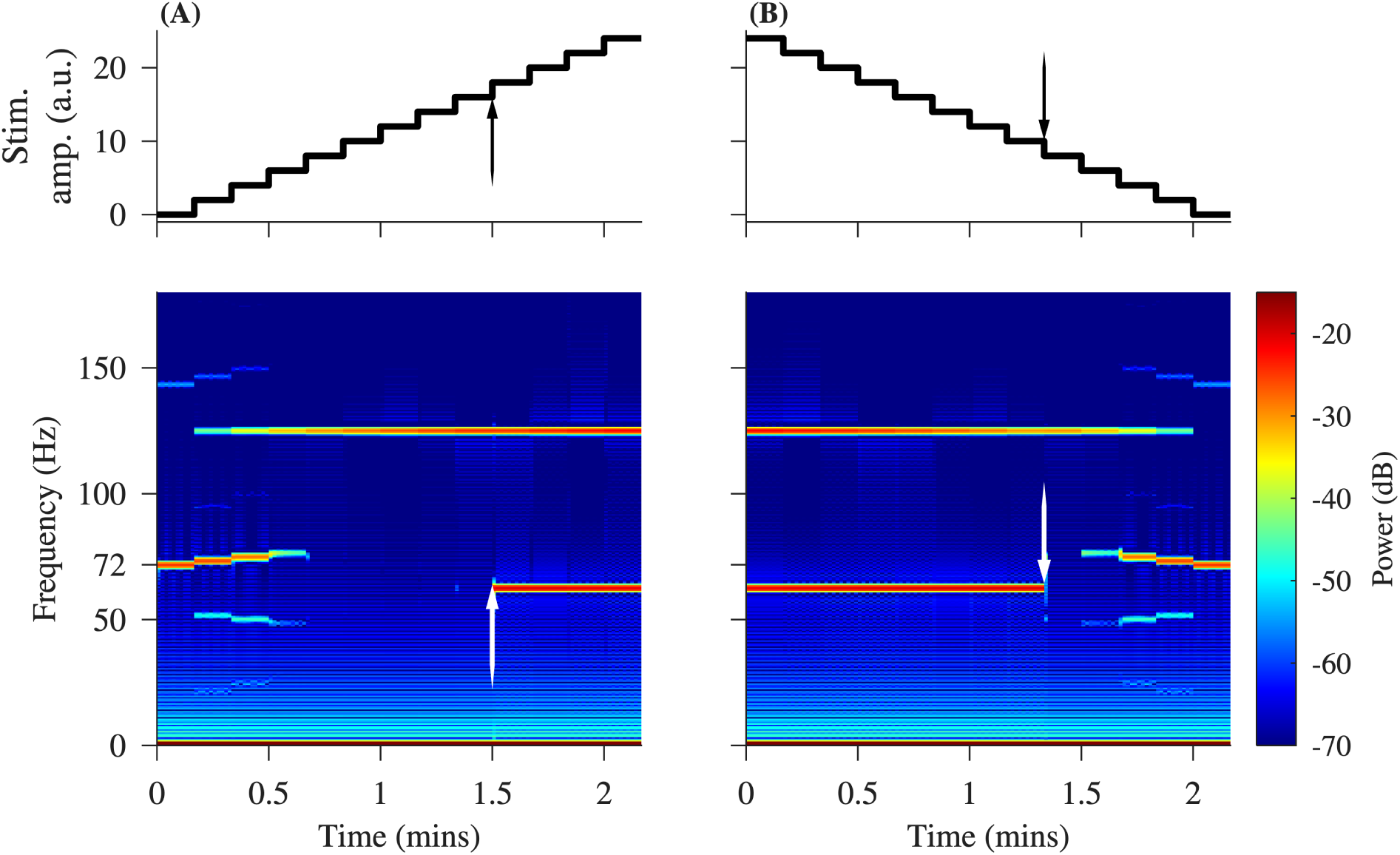
The effect of noise on simulated power spectral densities (PSD). Noise (right-hand panels) smears out the sharp peaks observed in the noise-free system (left-hand panels), but the qualitative features remain. (A) to (D) with strongly-damped oscillations (*ω_EE_* = 4.0, green curve) and self-sustained oscillations (*ω_EE_* = 5.2, red curve). (A) Without noise, stimulation off; (B) with noise, stimulation off; (C) without noise, stimulation on; (D) with noise, stimulation on. Stimulation amplitude, 4 a.u.; stimulation frequency, 125 Hz. All other parameters take their baseline values as given in the Supplementary Materials.

Specifically, in the absence of stimulation and without noise, see Fig. 9(A), in the strongly-damped case, there was no spectral power to machine precision; whereas, in the case of self-sustained oscillations there was a pronounced peak at around 70 Hz. For this value of *ω_EE_*, the system is sufficiently far from the Hopf bifurcation that the oscillation is not purely sinusoidal, as reflected by the additional low amplitude super-harmonic response centred at 140 Hz. Note that this super-harmonic peak was several orders of magnitude lower in amplitude that the harmonic component.

When noise was added, see Fig. 9(B), in the strongly-damped case, a broadband peak was observed due to noise-induced oscillations centred around the frequency of the damped oscillations. In the case of self-sustained oscillations, both the peak at the natural frequency of 70 Hz and its super-harmonic remain but are flattened and broadened out.

On application of DBS and without noise, see Fig. 9(C), in the strongly-damped case only the sharp peak at the stimulation frequency is visible. In the self-sustained oscillations case, the peak associated at the natural frequency shifted to 73 Hz, a subharmonic of the stimulation frequency with *ρ* = 0.58. In addition, there was a sharp peak at the stimulation frequency. When noise was added, see Fig. 9(D), peaks were in general broadened out and flattened.

The blurring out of sharp peaks was also reflected in a blurring of boundaries in the stimulation frequency-amplitude space, compare Fig. S6 with the deterministic stimulation parameter planes shown in Fig. S3, Fig. 4 and Fig. 5. In the strongly-damped case, a small region of responses other than *ρ* = 1 were seen for high-frequency low-amplitude stimulation (Fig. S6(A), left). These were due to noise-induced oscillations at a frequency close to the natural frequency of the damped oscillations. These solutions were very low in amplitude (Fig. S6(A), right).

The same blurring out of boundaries and broadening of peaks was also observed when simulation amplitude was ramped up, see Supplementary Materials Fig. S7. In the strongly-damped case, there was a sharp peak at the stimulation frequency as in the noise-free case (c.f. Fig. 6) but there was in addition evidence of the low amplitude noise-induced oscillations centred at the natural frequency of the damped oscillations, approximately 80 Hz. These broadband noise-induced oscillations drifted towards the half-harmonic response and became entrained to the half-harmonic stimulation frequency at high enough stimulation amplitudes above 16 a.u., see Supplementary Fig. S7.

### 3.7 Sensitivity analysis

Perturbing parameters around their baseline values yielded results as expected from the underlying dynamical structure.

Perturbations in parameters around the strongly-damped case, yielded no qualitative change to the responses in the frequency-amplitude stimulation plane. Oscillations remained strongly-damped and the only responses to stimulation were harmonic to the drive (*ρ* = 1).

For the weakly-damped case, perturbations from the baseline parameter values continued to lead to weakly-damped oscillations for all but one case (increasing *ω_EE_*). Parameter changes which took the system further away from the Hopf bifurcation led to an increase in the required stimulation amplitude for generating a half-harmonic response with a corresponding decrease in the area of the half-harmonic tongue, as illustrated for varying parameter *ω_EI_* in Supplementary Fig. S8. The changes in the amplitude of stimulation at the tip of the half-harmonic tongue, the area of the tongue and the frequency for the half-harmonic at the tip of the tongue for all 7 parameters are shown in Supplementary Fig. S9. On increasing *ω_EE_*by 5%, the system crosses the Hopf bifurcation point located at *ω_EE_* = 4.992 into the region of existence of self-sustained oscillations. Correspondingly, in the stimulation frequency-amplitude plane, the amplitude of stimulation required to generate a half-harmonic response was reduced to zero and many other subharmonic responses became possible.

For the self-sustained case, perturbations from the baseline parameter values continued to lead to self-sustained oscillations for all but one case. Applying stimulation then resulted in an Arnold tongue structure with multiple subharmonic tongues. On decreasing *ω_EE_* by 5%, the system crosses the Hopf bifurcation point located at *ω_EE_* = 4.992 into the region of weakly-damped oscillations. Summary metrics describing how the position and area of the half-harmonic tongue change for all 7 parameters are presented in Supplementary Fig. S9. An example illustrating how the tongue structure changes for varying parameter *τ_I_* is shown in Supplementary Fig. S10.

For each case, we investigated the presence of hysteresis. When the resulting unforced system had self-sustained oscillations, the hysteresis region persisted and its area and shape varied with parameters. In some cases, the area of hysteresis overlapped with the region of complex dynamics.

An overview of the qualitative changes in the dynamical regimes as a result of individual parameter perturbations is summarised in Supplementary Table S4.

## 4 Discussion

FTG rhythms have been observed in multiple clinical studies. In most patients FTG rhythms have been shown to have prokinetic effects, but there is some evidence they are associated with dyskinesia, particularly when no sFTG is found off-stimulation [30, 18]. In addition, FTG has been proposed as a putative biomarker for adaptive DBS systems [36]. Whether to increase FTG rhythms to enhance prokinetic effects, suppress FTG rhythms to reduce dyskinesia or use FTG rhythms as a biomarker, there is a need for theoretical frameworks to aid understanding.

### 4.1 Broad theoretical framework and relation to finely tuned gamma (FTG) rhythms

Here, we have set the previously-discussed entrainment of gamma oscillations in PD in a broader theoretical framework.

Instead of referring to Arnold tongues as the explanation for all half-harmonic enhancement of gamma oscillations, here we argue that patients who exhibit no sFTG off-stimulation and no half-harmonic response on stimulation are consistent with physiological parameter regimes in which there are only strongly-damped gamma oscillations.

Patients who exhibit no sFTG off-stimulation and half-harmonic responses on stimulation agree with physiological parameter regimes in which there are weakly-damped gamma oscillations. This challenges the previous assumption that all patients exhibiting a half-harmonic response to the stimulation must also exhibit sFTG, even if the oscillation is too small to be detected in the LFP measurements [30, 42, 32].

Patients who exhibit sFTG off-stimulation which are entrained to give a half-harmonic response on stimulation can be modelled by physiological parameter regimes in which there are self-sustained gamma oscillations.

### 4.2 Consequences of the theoretical framework

This broad theoretical framework highlights three generic consequences. First, in the absence of self-sustained oscillations (no-sFTG), the model predicts that there is no self-sustained rhythm to entrain, and the system is more accurately described as a driven system. Driven systems typically respond at frequencies which are harmonic to the stimulation (drive). To respond at a half-harmonic the system needs to be nonlinear, and the stimulation has to have a sufficiently large amplitude. Stimulation amplitude has to be sufficiently large because, generically, even in nonlinear systems, in the neighbourhood of equilibrium solutions in phase space the dynamics are linear. Consequently, stimulation needs to be sufficiently large to perturb the system out of this local linear neighbourhood.

Second, the mathematical formulation suggests that in the presence of self-sustained oscillations (sFTG), oscillations can be entrained at frequencies which are harmonic or at multiple different subharmonics to the stimulation. We note that our results highlight that the three simulated half-harmonic tongue-like regions shown in [41] are not Arnold tongues in the conventional sense, since for the parameter values used there were no self-sustained oscillation to entrain. Within our wider theoretical framework, these tongue-like regions are a driven nonlinear response.

We have not found any published reports of subharmonics other than the half-harmonic, but we are aware that observations of other subharmonic responses may have been underreported in the past. In Fig. S11, we show example patient data for the case of a *ρ* = 1*/*4 subharmonic in which an endogenous rhythm of approximately 50 Hz is entrained to one quarter of the stimulation frequency for some stimulation amplitudes. These data were collected by some of the co-authors and analysed using the approaches specified in [8].

Third, characteristic of nonlinear systems, we hypothesise that there may be regions in stimulation parameter space where there is co-existence of solutions i.e. the same stimulation parameters lead to different responses depending on the initial conditions (i.e. past history). Consequently, changes in stimulation may result with hysteresis in responses. Hysteresis in biological and physiological phenomena has been observed and modelled in other systems, including stimulation of ventricular cells [55] or entrainment of circadian rhythms by multiple zeitgebers [35]. Such path-dependent responses pose a challenge for adaptive stimulation algorithms since a simple feedback-control system will have difficulty tracking and controlling responses that do not vary continuously and monotonically with control parameters. Hysteresis has not yet been reported, to our knowledge, in the case of neuromodulation.

### 4.3 Robustness of the theoretical framework

The results we have present are based on a minimal physiological model and general dynamical systems theory which indicate that response to forcing (stimulation) should differ substantially between damped systems and those with self-sustained oscillations [20, 15].

We provided a sketch proof to show that low-dimensional linear driven systems with impulsive forcing cannot exhibit subharmonic response. Although we only considered impulsive stimulation and low-dimensional systems, the general principles behind dynamical systems argue that sufficiently close to stable equilibria, behaviour is dominated by the slowest decaying modes and may be approximated by a local linear system. Since half-harmonic responses are observed, the implications are that, first, not surprisingly, the dynamics of the brain are nonlinear. Second, for half-harmonic responses to occur when oscillations are damped, the stimulation has to be sufficiently strong for nonlinearity to have an impact.

The Wilson-Cowan model includes seven physiological parameters. We only considered varying three of them (*ω_EE_, ω_EI_, ω_IE_*). As other parameters are varied, the number of equilibria and the regions of oscillatory behaviour will change. Indeed, in the Supplementary Materials we demonstrate that up to five equilibria can occur depending on the parameter values. However, the central theme that response to stimulation will depend on whether oscillations are strongly-damped, weakly-damped or self-sustained relies on generic properties of dynamical systems which are agnostic to the details.

In a noise-free model, the transition between damped and self-sustained oscillations occurs at Hopf bifurcations at well-defined parameter values. But in the presence of noise, more representative of a physiological system, the distinction is blurred.

In our simulations, following previous work [41], only additive Gaussian noise was used as a minimal representation of background fluctuations and measurement error. Given that our central results arise from the underlying nature of the deterministic system and the presence (or absence) of self-sustained oscillations we anticipate that a moderate-coloured noise might change the shape or incline of the enhancement tongues but will not fundamentally alter our conclusions.

Sensitivity analysis revealed that the behaviour of the tongues changed in a manner that was consistent with the broader theoretical framework. Hysteresis was consistently found when self-sustained oscillations were present off-stimulation.

We note that the general dynamical framework we set out is agnostic to brain region and is not specific to gamma rhythms. In patients with Parkinson’s Disease, half-harmonic responses have been reported from recordings in the cortex, STN and GPi. Recent work has revealed half-harmonic responses in the gamma band with stimulation of the ventral intermediate nucleus in patients with Multiple Sclerosis [38]. Also consistent with our framework is harmonic entrainment of sFTG by applying stimulation in the gamma band, at the frequency of recorded sFTG [49].

### 4.4 Other theories

An alternative explanation for the appearance of half-harmonic responses is a priming-like mechanism. In the context of DBS, priming has been investigated in the form of delivering paired pulses and observing that the response to the second pulse differed from the first one [4, 6]. While this could be a basis for the gamma responses, we are not aware of any reports showing that paired pulses directly result in half-harmonic oscillations in continuous DBS delivery. Priming and the entrainment frameworks of modelling sub-harmonic responses should not be considered as contradictory views. While examples of priming, also in clinical context, explain the mechanism at a single-neuron level, the presented Wilson-Cowan model captures the behaviour at the neuronal population level.

### 4.5 Limitations

Our results rely on broad principles of driven nonlinear dynamical systems and a minimal physiological model. However, the phenomenological nature of the model is a limitation in that it is not possible to make direct comparison between model parameters and physiology, and stimulation amplitudes are necessarily in arbitrary units. The presented form of the model was initially developed in [41], with parameters fitted and model predictions verified in patients with both STN and GPi implanted DBS. In all these cases, recordings were taken using ECoG arrays, suggesting that the model captures general features and phenomena relating to DBS across different stimulation and recording locations. However, this does not imply that stimulation at STN and GPi have an equivalent physiological outcome. In fact, response to stimulation differs depending on the target location on micro-, meso-, and macro-scales [34]. STN and GPi engage different upstream and downstream patterns, with different levels and balances of excitatory and inhibitory inputs, which could influence the parameters of the model. However, sufficiently robust and large-scale studies with structured changes to stimulation regimes at different targets are currently lacking. Limiting our ability to accurately comment and hypothesise about the link between parameter choices and stimulation and recording location. Given the physiological complexity and the lack of consensus on the mechanism by which DBS acts, we are some way from developing a spatially-resolved model which includes all relevant networks and stimulation effects. It is in this context that our minimal model is of value as it captures broad features and highlights classes of phenomena which should be expected to occur.

In addition, we have only focussed on oscillations in the gamma band. The interplay between other prominent rhythms observed in PD, in particular, the bursts of beta rhythms and their suppression, may prove crucial in fully understanding the mechanisms behind DBS.

The mechanisms described by this framework do not capture DBS-induced FTG, observed in patients off-medication. In [52], STN stimulation in the absence of levodopa induced gamma oscillations but found no consistent response of these oscillations to changes in stimulation frequency. Furthermore, oscillations induced in that way persisted after the stimulation was terminated, further arguing against a direct entrainment mechanism. This strongly suggests that the structure of the network and the properties of the model are strongly affected by the medication state. The two mechanisms should not be seen as mutually exclusive – our framework highlights that the presence of weakly-damped or self-sustained oscillations to drive or entrain the rhythm to a stimulation frequency-dependent response. Capturing DBS-induced FTG together with its long transient after stimulation is switched off, is likely to require the networks dynamics to be described at a different, slower timescale. This is not included in the current framework.

Finally, even in this minimal model we found regions of complex dynamics at high stimulation amplitudes, which we have not fully investigated. This includes period-doubling and chaotic dynamics characteristic of other forced systems [16, 21], although in the presence of noise these more complex dynamics may not be observable.

### 4.6 Final words and further extensions

We have assumed that physiological parameter values are fixed. In reality, the balance between excitation and inhibition is likely to change depending on activity, sleep/wake state, or ultradian and circadian oscillations [45, 56, 50]. As physiological parameters vary, as shown in Fig. 3, damped oscillations may change to self-sustained oscillations or vice-versa. In addition, damped oscillations may change from being weakly damped (near Hopf bifurcations) or strongly damped. Hence making parameter values medication-dependent and / or brain state (sleep/wake) dependent and / or time-of-day dependent may explain how gamma band responses change over time.

Overall, our results show that the Wilson-Cowan framework offers a possible dynamical systems explanation of the considerable heterogeneity in responses of oscillations in the gamma range to DBS and highlights that subharmonic responses, not only harmonic and half-harmonic responses, are a predicted feature of entrained systems. Our modelling framework could guide further experimental studies to better understand the complex brain–stimulus interactions to improve adaptive DBS.

## Data Availability

No new data has been used in this study.

## Acknowledgements

The authors would like to thank Timothy Constandinou for helpful conversations and his input into the project.

## CRediT authorship contribution statement

**Stanisław W. Biber:** conceptualization, investigation, methodology, visualization, writing – original draft, writing – review and editing; **James J. Sermon:** conceptualization; **Jonathan Kaplan:** writing – review and editing; **Johannes L. Busch:** writing – review and editing; **Andrea A. Kühn:** writing – review and editing; **Derk-Jan Dijk:** conceptualization, funding acquisition, writing – review and editing; **Timothy Denison:** conceptualization, funding acquisition, writing – review and editing; **Anne C. Skeldon:** conceptualization, funding acquisition, methodology, supervision, writing – original draft, writing – review and editing.

