## Supplementary Materials for "Heterogeneity in deep brain stimulation induced gamma oscillations may be explained by bifurcations in neural dynamics"

S.W. Biber, J.J. Sermon, J. Kaplan, J.L. Busch, A.A. Kühn, D.-J. Dijk, T. Denison,

A.C. Skeldon

### 1 Model equations & analysis of the equilibria

#### 1.1 Model equations

We consider a model of stimulation based on the Wilson-Cowan modelling framework [3, 6].

Formally, the model is described as

$$\begin{aligned} \mathrm{d}E &= \frac{1}{\tau_E}\left[-E + f_b\left(\eta_E + \omega_{EE}E - \omega_{EI}I\right)\right]\mathrm{d}t + \zeta \mathrm{d}W_E, \\ \mathrm{d}I &= \frac{1}{\tau_I}\left[-I + f_b\left(-\eta_I + \omega_{IE}E\right) + A(t)\right]\mathrm{d}t + \zeta \mathrm{d}W_I, \end{aligned} \tag{S1}$$

where $f_b(x) = [1 + \exp(-b(x-1))]^{-1}$ is a switch-like activation function. For low values of the argument $x$, $f_b(x)$ asymptotes to 0, and for high values $f_b(x)$ asymptotes to 1. This switch-like function models the fact that inputs to neuronal populations have to be sufficiently large to have an effect. $A(t)$ is the DBS forcing term. Stochastic effects are represented by Wiener processes $\mathrm{d}W_{E,I}$, with standard deviation $\zeta$ (assumed to be the same for both neuronal populations). We consider the behaviour of the system both without noise ($\zeta = 0$) and with noise ($\zeta > 0$).

#### 1.2 Analysis of equilibria

Here, we consider the deterministic version of the system given by Equations (S1), that is, taking $\zeta = 0$.

We consider the nature of the system equilibria, that is we consider points $(E^*, I^*)$ such that $\frac{\mathrm{d}E}{\mathrm{d}t}\big|_{(E^*,I^*)} = 0 = \frac{\mathrm{d}I}{\mathrm{d}t}\big|_{(E^*,I^*)}$.

At any equilibrium $(E^*, I^*)$, the Jacobian matrix of the system becomes

$$J = \begin{bmatrix} \frac{1}{\tau_E}\left(b\omega_{EE}\, E_Q - 1\right) & -\frac{1}{\tau_E}\left(b\omega_{IE}\, E_Q\right) \\ \frac{1}{\tau_I}\left(b\omega_{EI}\, I_Q\right) & -\frac{1}{\tau_I} \end{bmatrix}, \tag{S2}$$

where $E_Q = E^*(1-E^*)$, $I_Q = I^*(1-I^*)$.

The eigenvalues of the linearised system are therefore

$$\begin{aligned} \lambda_{\pm} = & \frac{(b\omega_{EE}\, E_Q - 1)\tau_I - \tau_E}{2\tau_E\tau_I} \\ & \pm \frac{\left[b^2\omega_{EE}^2\, E_Q^2\tau_I^2 + (\tau_E - \tau_I)^2 - 2b\omega_{EE}\, E_Q(\tau_I^2 - \tau_E\tau_I) - 4b^2\omega_{IE}\omega_{EI}\, E_Q\, I_Q\tau_E\tau_I\right]^{1/2}}{2\tau_E\tau_I}. \end{aligned} \tag{S3}$$

At a Hopf bifurcation, one requires $\mathrm{Re}(\lambda) = 0$, yielding

$$E_Q = \frac{\frac{\tau_E}{\tau_I} + 1}{b\omega_{EE}}, \tag{S4}$$

and hence

$$E^* = \frac{1 \pm \left[1 - \frac{4(T+1)}{b\omega_{EE}}\right]^{1/2}}{2}, \tag{S5}$$

where $T = \frac{\tau_E}{\tau_I}$, and only solutions $0 < E^* < 1$ are admitted.

A further simplification also gives

$$\mathrm{Im}(\lambda) = \frac{1}{2\tau_E\tau_I}\left[4b\frac{\omega_{IE}\omega_{EI}}{\omega_{EE}}(\tau_E\tau_I + \tau_E^2)\, I_Q - 4\tau_E^2\right]^{1/2}. \tag{S6}$$

This, in turn requires that

$$I_Q > \frac{\tau_E}{b(\tau_I + \tau_E)}\frac{\omega_{EE}}{\omega_{IE}\omega_{EI}}. \tag{S7}$$

Combining together, to solve for a parameter value at which Hopf bifurcation occurs, it is required to solve

$$\begin{aligned} E^* &= f_b\left(\eta_E + \omega_{EE}E^* - \omega_{EI}f_b\left(-\eta_I + \omega_{IE}E\right)\right) \\ E^* &= \frac{1 \pm \left[1 - \frac{4(\tau_E/\tau_I+1)}{b\omega_{EE}}\right]^{1/2}}{2}, \end{aligned} \tag{S8}$$

subject to the solutions $E^*, I^* = f_b\left(-\eta_I + \omega_{IE}E^*\right)$ and the chosen parameter satisfying the condition (S7).

The nullclines of the system for different parameters, demonstrating the existence of one, three or five equilibria, are given in Fig. S1.

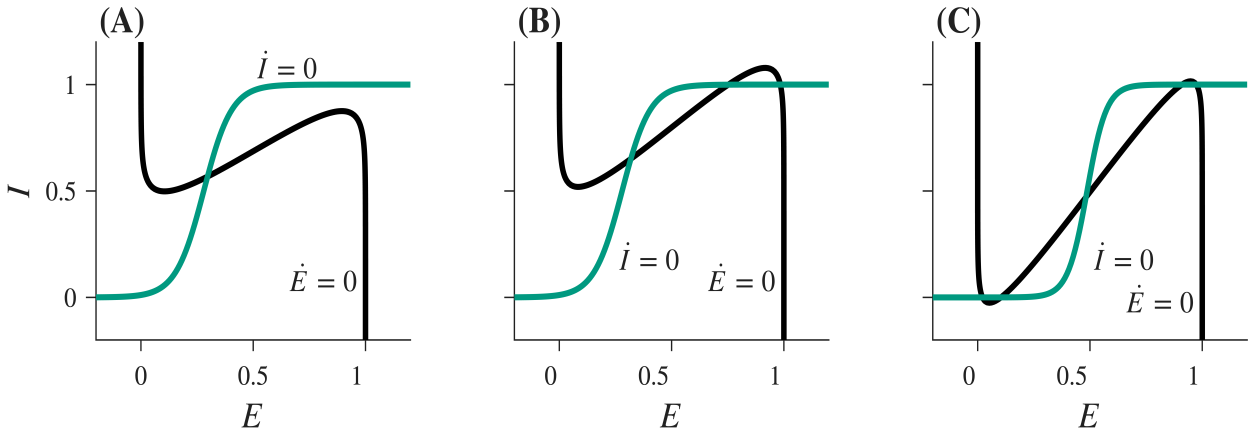


Figure S1: Nullclines for system (S1). Parameters are as presented in Table S3 with the following changes: (A) $\omega_{EE} = 4.795$, (B) $\omega_{EE} = 5.895$, (C) $\omega_{EE} = 6.295, \omega_{IE} = 3.7, \eta_E = -0.323, \eta_I = -2.52, b = 3.22$.

#### 2 Table of abbreviations

Table S1: Table of abbreviations and terminology used throughout the paper.

| Abbreviation | Meaning | Physiological description (if applicable) | Mathematical description (if applicable) |
|---|---|---|---|
| a.u. | Arbitrary units | — | Dimensionless amplitude unit used for the stimulation input. |
| DBS | Deep brain stimulation | — | Modelled as an external forcing term $A(t)$. |
| FTG | Finely tuned gamma | Narrowband gamma-band oscillatory activity. | Stable oscillatory behaviour. |
| GPi | Globus pallidus internus | — | — |
| LFP | Local field potential | — | — |
| PD | Parkinson's disease | — | — |
| PSD | Power spectral density | — | — |
| sFTG | Spontaneous finely tuned gamma | Endogenous, narrowband gamma activity observed without stimulation, particularly in some patients receiving dopaminergic medication. | The self-sustained oscillation observed in the absence of forcing. |
| STN | Subthalamic nucleus | — | — |

### 3 Parameter values used in simulations

Table S3 presents the parameter values for the Wilson-Cowan model (eq. (S1)) are taken from participant RCS02 from [4].

In our simulations, we alter the value of the self-excitation parameter $\omega_{EE}$. We chose three particular cases, which reflect different natures of the system.

For the case of no self-sustained oscillation, and where the half-harmonic oscillation cannot be induced by the stimulation, we take $\omega_{EE} = 4.0$.

For the case of no self-sustained oscillation, but where the half-harmonic oscillation is induced by the stimulation, we take $\omega_{EE} = 4.795$, identical to the estimate from [4].

For the case with self-sustained oscillation, we take $\omega_{EE} = 5.2$.

Table S3: Parameter values used in numerical simulations of the Wilson-Cowan model.

| $\omega_{EI}$ | $\omega_{IE}$ | $\tau_E$ | $\tau_I$ | $\eta_E$ | $\eta_I$ | b | $\zeta$ |
|---|---|---|---|---|---|---|---|
| 7.224 | 4.913 | $6.424 \times 10^{-3}$ | $4.853 \times 10^{-3}$ | 1.977 | $-1.020$ | 2.216 | 0.087 |

### 4 Sub-harmonic responses in weakly-damped systems

#### 4.1 Nonlinearity as a necessary condition for half-harmonic responses

In the main text, we have shown that a linear ODE system with a stable equilibrium may not give rise to a half-harmonic oscillation with regard to an instantaneous periodic forcing.

Here, we show that the proof extends to 2-dimensional linear systems. In the case of systems with a stable node, the proof is a simple extension of the 1-dimensional case. Considering a system with a stable spiral requires additional consideration. In all cases, the strategy of the proof is the same and goes as follows. We assume the system to start from a set point and apply two stimulation pulses at equal time intervals $T$. After two periods ($2T$) we assume the system returns to its original position (rotation number $\rho = 1/2$) and show that this is only possible if it previously returned to that position at time $T$ (harmonic entrainment).

We consider a planar system

$$\dot{\boldsymbol{x}} = A\boldsymbol{x}, \tag{S9}$$

where $\boldsymbol{x} \in \mathbb{R}^2$, and $A \in \mathbb{R}^{2\times 2}$.

Suppose $A$ has two distinct negative eigenvalues $-\lambda_1, -\lambda_2$, with eigenvectors $\boldsymbol{v}_1$ and $\boldsymbol{v}_2$. Let $\boldsymbol{w} = V\boldsymbol{x}$, where $V = [\boldsymbol{v}_1, \boldsymbol{v}_2]^{-1}$ (that is, we redefine the system in the eigenbasis). Then, the system can be rewritten as

$$\begin{aligned} \dot{w}_1(t) &= -\lambda_1 w_1(t), \\ \dot{w}_2(t) &= -\lambda_2 w_2(t), \end{aligned} \tag{S10}$$

which is equivalent to two 1-dimensional systems, and hence the same argument as before follows. This also holds if $\lambda_1 = \lambda_2$, but the geometric multiplicity of this eigenvalue is two.

Now, consider the case where $-\lambda_1 = -\lambda_2 = -\lambda$, that is, the eigenvalue has the algebraic multiplicity of two, but we focus on the case where its geometric multiplicity is one.

Once again, we redefine the variables in the eigenbasis $\boldsymbol{w} = V\boldsymbol{x}$, where $V$ is now a matrix of Jordan eigenvectors. The system can now be written as

$$\begin{aligned} \dot{w}_1(t) &= -\lambda w_1(t) + w_2(t), \\ \dot{w}_2(t) &= -\lambda w_2(t), \end{aligned} \tag{S11}$$

yielding solutions of a form

$$\begin{aligned} w_1(t) &= A\mathrm{e}^{-\lambda t} + Bt\mathrm{e}^{-\lambda t}, \\ w_2(t) &= B\mathrm{e}^{-\lambda t}. \end{aligned} \tag{S12}$$

Suppose the starting point for the system is now $\boldsymbol{w}_0 = [\varepsilon_1, \varepsilon_2]^\intercal$, and the pulse is now $\Delta = [\Delta_1, \Delta_2]^\intercal$. For $w_2(t)$, the same argument follows as before; hence, we know only harmonic oscillation is possible, and in particular $w_2(T) = (\varepsilon_2 + \Delta_2)\mathrm{e}^{-\lambda T} = \varepsilon_2$ before the pulse is applied for the second time at $t = T$.

For $w_1(t)$, after the first stimulation (that is, for $t \in (0, T)$) the solution takes the form $w_1(t) = (\varepsilon_1 + \Delta_1)\mathrm{e}^{-\lambda t} + t(\varepsilon_2 + \Delta_2)\mathrm{e}^{-\lambda t}$. After the second impulse (that is $t \in (T, 2T)$), by recalculating the integration constants, we have that $w_1(t) = \left[\varepsilon_1 + \Delta_1 + (\Delta_1 - \Delta_2 T)\mathrm{e}^{\lambda T}\right] \mathrm{e}^{-\lambda T} + t\left[(\varepsilon_2 + \Delta_2) + \Delta_2 \mathrm{e}^{\lambda T}\right] \mathrm{e}^{-\lambda t}$.

Requiring that $w_1(2T) = \varepsilon_1$ (which is necessary for the half-harmonic response), we again solve for $\mathrm{e}^{-\lambda T}$, resulting with $\mathrm{e}^{-\lambda T} = \frac{\varepsilon_1 - T\varepsilon_2}{\Delta_1 + \varepsilon_1}$. This, in turn, yields

$$\begin{aligned} w_1(T) &= (\varepsilon_1 + \Delta_1)\mathrm{e}^{-\lambda T} + T(\varepsilon_2 + \Delta_2)\mathrm{e}^{-\lambda T} \\ &= (\varepsilon_1 + \Delta_1)\frac{\varepsilon_1 - T\varepsilon_2}{\Delta_1 + \varepsilon_1} + T\varepsilon_2 = \varepsilon_1, \end{aligned} \tag{S13}$$

where we have also used $(\varepsilon_2 + \Delta_2)\mathrm{e}^{-\lambda T} = \varepsilon_2$, which follows from the harmonic nature of $w_2(t)$. This demonstrates that $w_1(t)$ is also entrained with $\rho = 1$.

Finally, consider the case of a stable spiral. That is, let $A$ have complex eigenvalues $\lambda = -\mu \pm i\omega$, where $\mu > 0$, with the corresponding eigenvectors $\boldsymbol{v} = \boldsymbol{p} \pm i\boldsymbol{q}$. Once again, by considering the solution in the eigenbasis, we define $\boldsymbol{w} = V\boldsymbol{x}$, where $V = [\boldsymbol{p}, \boldsymbol{q}]^{-1}$. In the new variables, our system has the solution

$$\begin{aligned} w_1(t) &= \mathrm{e}^{-\mu t}\left[C_1 \cos\omega t + C_2 \sin\omega t\right], \\ w_2(t) &= \mathrm{e}^{-\mu t}\left[C_2 \cos\omega t - C_1 \sin\omega t\right]. \end{aligned} \tag{S14}$$

Again, let the system start at $t = 0$ at $\boldsymbol{w}_0 = [\varepsilon_1, \varepsilon_2]^\intercal$ and apply an instantaneous pulse $\Delta = [\Delta_1, \Delta_2]^\intercal$.

After the first pulse, the solutions are

$$\begin{aligned} w_1(t) &= \mathrm{e}^{-\mu t}\left[(\varepsilon_1 + \Delta_1)\cos\omega t + (\varepsilon_2 + \Delta_2)\sin\omega t\right], \\ w_2(t) &= \mathrm{e}^{-\mu t}\left[(\varepsilon_2 + \Delta_2)\cos\omega t - (\varepsilon_1 + \Delta_1)\sin\omega t\right] \end{aligned} \tag{S15}$$

for $t \in (0, T)$, and in particular at $t = T$ let

$$\begin{aligned} w_1(T) &= \mathrm{e}^{-\mu T}\left[(\varepsilon_1 + \Delta_1)\cos\omega T + (\varepsilon_2 + \Delta_2)\sin\omega T\right] = W_1, \\ w_2(T) &= \mathrm{e}^{-\mu T}\left[(\varepsilon_2 + \Delta_2)\cos\omega T - (\varepsilon_1 + \Delta_1)\sin\omega T\right] = W_2. \end{aligned} \tag{S16}$$

We now apply the second instantaneous stimulation pulse, so that after it is applied $w_1(T) = W_1 + \Delta_1$, $w_2(T) = W_2 + \Delta_2$. Recalculating the integration constants, in the time interval $t \in (T, 2T)$ we have

$$\begin{aligned} w_1(t) &= \mathrm{e}^{-\mu t}\left[\left(\mathrm{e}^{\mu T}\left((\Delta_1 + W_1)\cos\omega T - (\Delta_2 + W_2)\sin\omega T\right)\right)\cos\omega t + \left(\mathrm{e}^{\mu T}\left((\Delta_1 + W_1)\sin\omega T + (\Delta_2 + W_2)\cos\omega T\right)\right)\sin\omega t\right], \\ w_2(t) &= \mathrm{e}^{-\mu t}\left[\left(\mathrm{e}^{\mu T}\left((\Delta_1 + W_1)\sin\omega T + (\Delta_2 + W_2)\cos\omega T\right)\right)\cos\omega t - \left(\mathrm{e}^{\mu T}\left((\Delta_1 + W_1)\cos\omega T - (\Delta_2 + W_2)\sin\omega T\right)\right)\sin\omega t\right]. \end{aligned} \tag{S17}$$

Evaluating now at the end of the second period (that is $t = 2T$) and requiring entrainment, yields

$$\begin{aligned} w_1(2T) &= \mathrm{e}^{-\mu T}\left[(W_1 + \Delta_1)\cos\omega T + (W_2 + \Delta_2)\sin\omega T\right] = \varepsilon_1, \\ w_2(2T) &= \mathrm{e}^{-\mu T}\left[(W_2 + \Delta_2)\cos\omega T - (W_1 + \Delta_1)\sin\omega T\right] = \varepsilon_2. \end{aligned} \tag{S18}$$

Noticing the symmetry with eq.(S16), we notice that it is only possible for those to be satisfied if $W_1 = \varepsilon_1$ and $W_2 = \varepsilon_2$, thus once again showing the necessity for the driven oscillation to be entrained at $\rho = 1$.

#### 4.2 Mechanism for 1:2 driven response – a geometric view

We illustrate a 1:2 driven response can occur in a nonlinear system using a geometric argument, contrasting it with the linear case.

We consider two trajectories, which are solutions to the same autonomous system, but starting at different initial conditions, see Fig. S2 (i). In the linear system, the distance between the two trajectories decreases monotonically with angular displacement (A). In a nonlinear system, the distance between the two trajectories does not necessarily decrease monotonically (B).

On application of a periodic sequence of stimulation pulses with a period $T$ which (instantaneously) increase the firing of the inhibitory population by $\Delta$, a 1:1 orbit is is possible in both linear and nonlinear systems, see Fig. S2(ii). In such a 1:1 orbit, the stimulation pulses connect a trajectory to itself in such a way that the change in $I$ is equal to $\Delta$ and the time taken for the system to evolve from the point on the phase plane corresponding to the end of one pulse to the point of the phase plane corresponding to the beginning of the next pulse is exactly the stimulation period. Changing the stimulation parameter (for example, changing the stimulation amplitude $\Delta$ or stimulation frequency) a 1:1 orbit continues to exist, although the precise orbit changes.

Now, constructing a 1:2 orbit requires connecting two trajectories, as illustrated in S2(A)(iii), such that: (i) the stimulation changes $I$ by $\Delta$; (ii) the time to evolve from one pulse to the next is exactly $T$, and (iii) that the pulses alternately take the system between two distinct orbits.

We demonstrated algebraically that it is not possible to construct a half-harmonic orbit in a linear system. In Fig. S2(A)(iii) we show that it is possible to satisfy the connectivity condition (i). However, it is not possible to simulataneously satisfy the requirement that the time between pulses are equal. Mathematically, this was demonstrated by the fact that evolution over two individual time intervals $T$ was equivalent to repeating the same trajectory twice. We see this in the symmetry of equations (S16) and (S18), where the exponential decrease of the spiral's radius $\mathrm{e}^{-\mu t}$ is applied twice and is equivalent to repeating the trajectory twice: $\left(\mathrm{e}^{-\mu T}\right)^2 = \mathrm{e}^{-2\mu T}$.

In the nonlinear case, the spiral's radius decreases as $\mathrm{e}^{-g(t)}$, for some nonlinear function $g(t)$, such that $\lim_{t\to\infty} g(t) = \infty$. The nonlinear nature of the function $g(t)$ means that the separation of the trajectories changes nonlinearly and may locally decrease and increase. The changing distance between the orbits allows for the two trajectories to be separated by the required distance at equal time intervals. Mathematically, this is expressed by the fact that evaluation over the time interval of $2T$ is not equivalent to two separate time intervals $T$, that is, $\left(\mathrm{e}^{-g(T)}\right)^2 = \mathrm{e}^{-2g(T)} \neq \mathrm{e}^{-g(2T)}$. This, in turn, arises from the nonlinear nature of the function $g(t)$, where $g(2T) \neq 2g(T)$.

Further work is required to explain why other sub-harmonics are not seen. For example, in this geometric interpretation, a 1:3 subharmonic could be thought of as requiring three trajectories connected by pulses of size $\Delta$ and separated by equal time intervals.

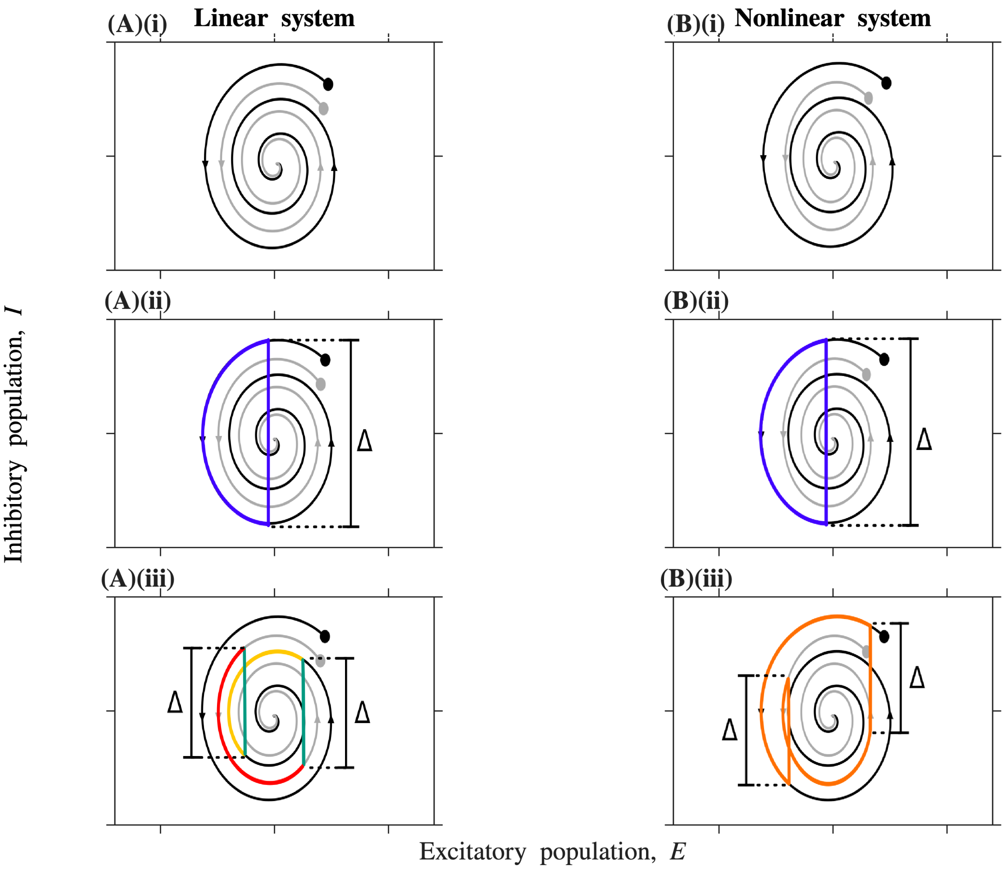


Figure S2: **Construction of 1:2 response orbit in the weakly-damped regime.** In all panels, black and grey trajectories correspond to distinct initial conditions, denoted with the '•' signs. Left panels (A) show the trajectories of a damped linear system. Right panels (B) show trajectories of a damped nonlinear system. Top panels (i) show the unforced trajectories. In the middle panel (ii) the blue trajectory shows a possible 1:1 orbit, where an unforced trajectory is linked to itself by the forcing. In the bottom panels (ii) 1:2 orbit is constructed for the nonlinear case by joining two distinct trajectories. In the solution to the linear system, it is possible to find two points of equal distance separation (green lines), but with an uneven time separation (red and orange curves).

#### 5 Supplementary figures & tables

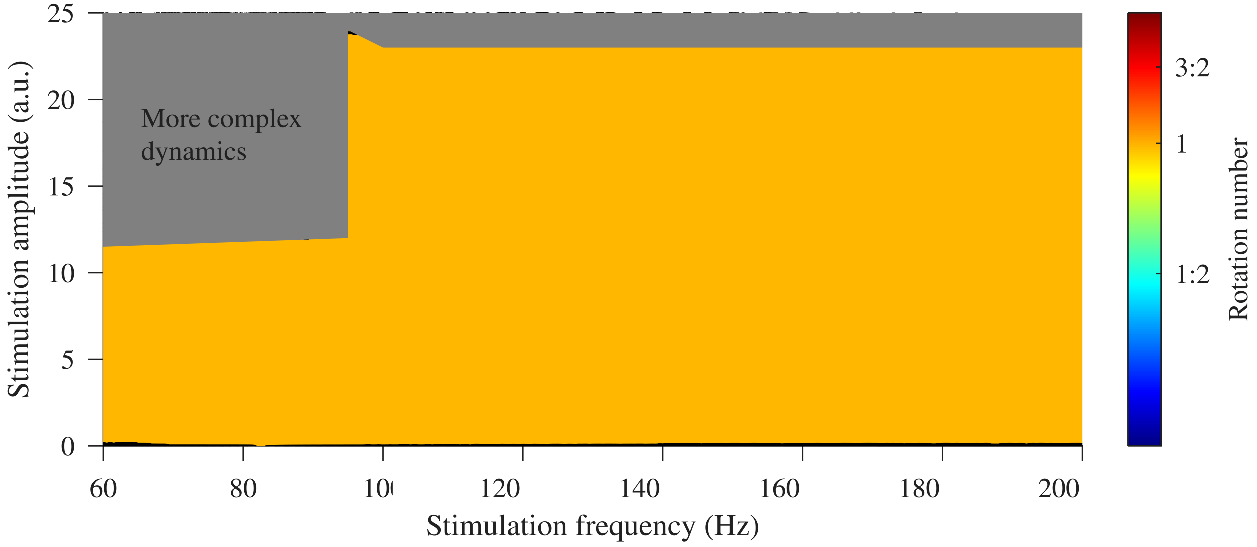


Figure S3: **Oscillatory behaviour as a function of stimulation amplitude and stimulation frequency in the presence of strongly-damped oscillations.** Stimulation parameter values resulting in harmonic responses are in orange, which covers the whole studied space. We observe more complex dynamics for high stimulation amplitude, which we do not discuss here.

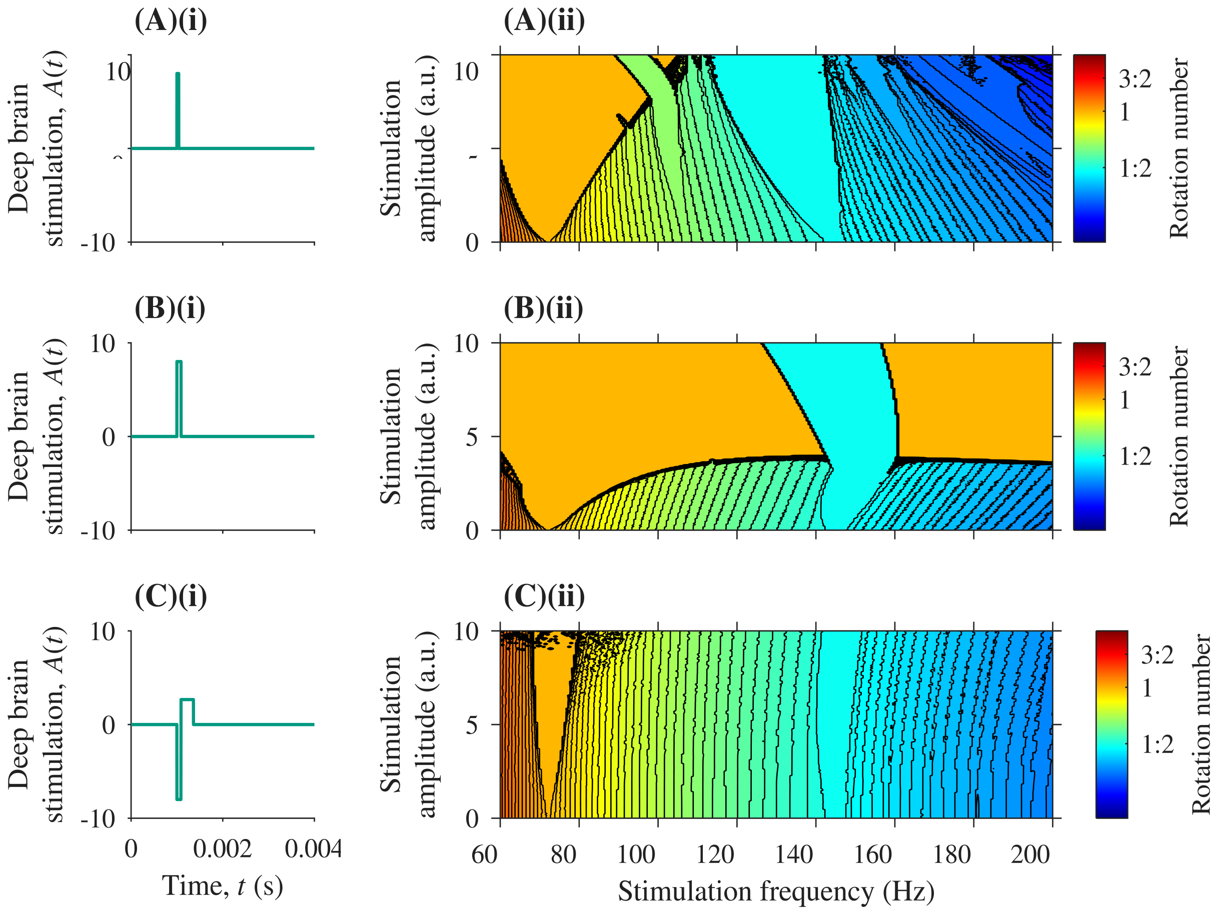


Figure S4: **Oscillatory behaviour as a function of stimulation amplitude and stimulation frequency for different stimulation waveforms.** Panels (i) display the waveform used, panels (ii) display the resulting entrainment structure. In (A), a narrow pulse is used (pulse width $4 \times 10^{-5}$ s). In (B), a wide pulse is used (pulse width $9 \times 10^{-5}$s). In (C), a biphasic pulse integrating to zero is used. Harmonic response ($\rho = 1$, orange), and half-harmonic responses ($\rho = 1/2$, cyan) are the largest regions.

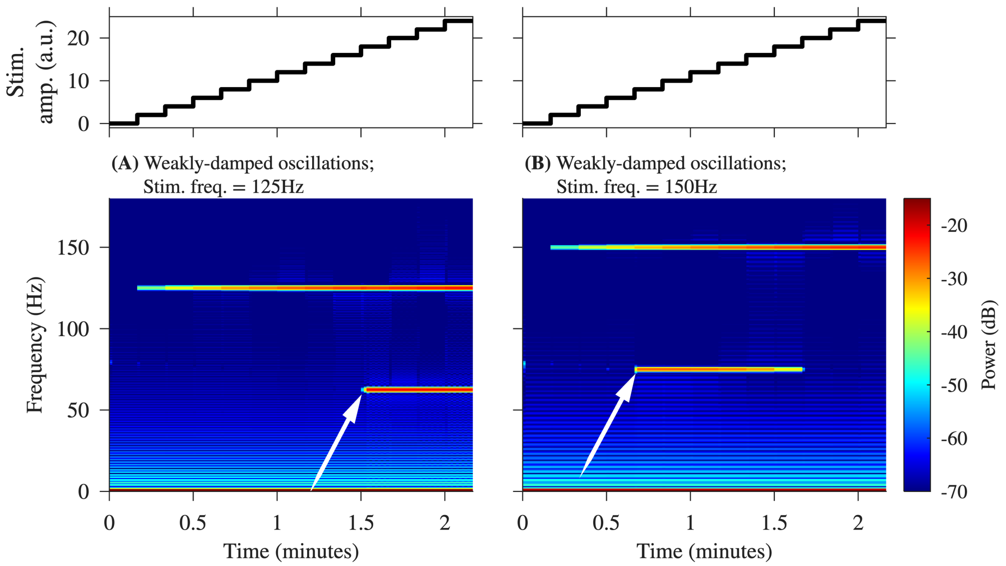


Figure S5: **Example predictions of a neuromodulation experiment.** Top panels show the time course of the stimulation amplitude as it is ramped up in discrete steps every 10 seconds. Spectrograms for two scenarios. Panel (A): stimulation frequency 125 Hz. Panel (B): stimulation frequency 150 Hz. Simulations of the system with weakly-damped oscillations ($\omega_{EE} = 4.795$). Baseline physiological parameters are given in the Supplementary Materials.

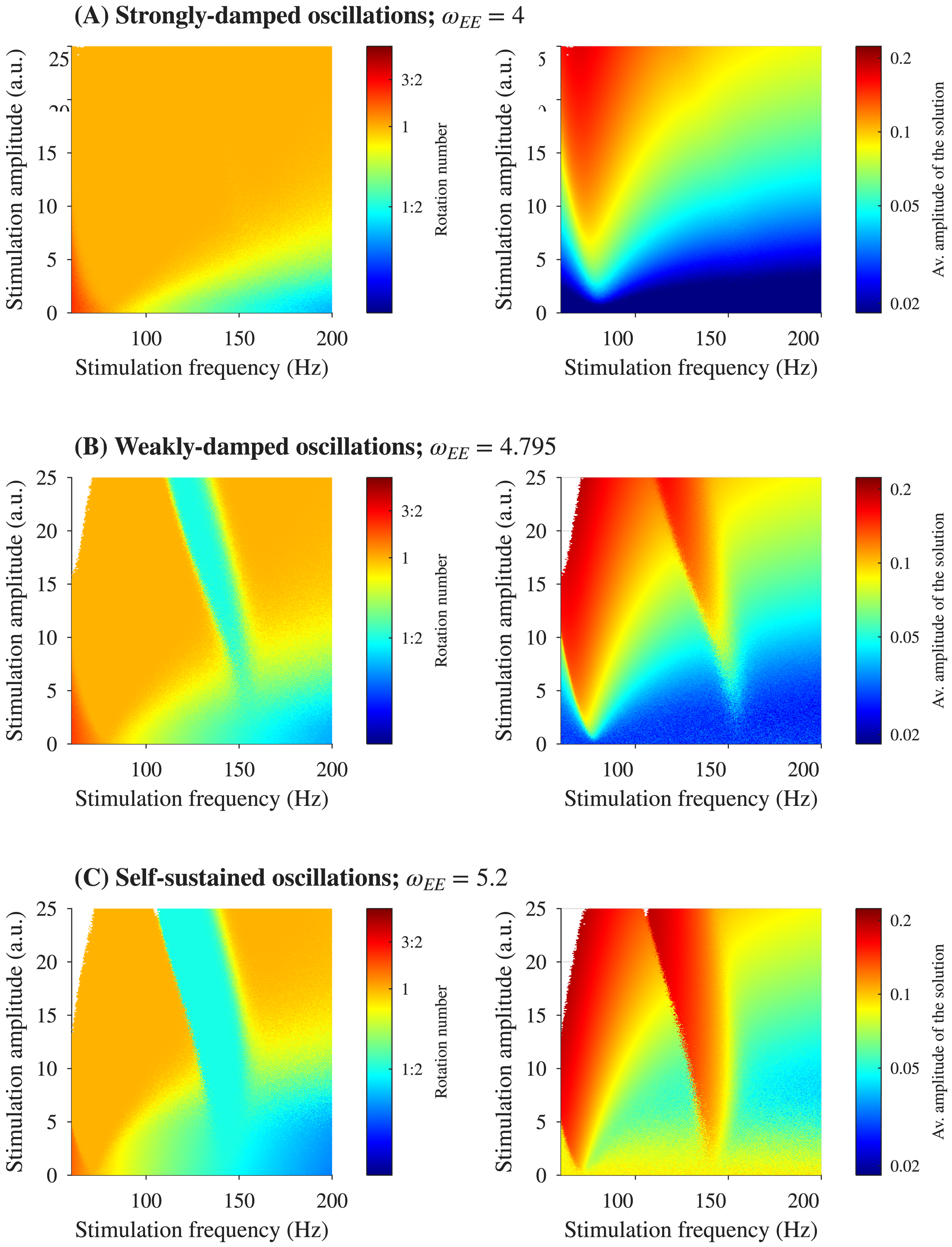


Figure S6: **Oscillatory behaviour as a function of stimulation amplitude and stimulation frequency in the presence of background noise.** (A) strongly-damped oscillations ($\omega_{EE} = 4.0$); (B) weakly-damped oscillations ($\omega_{EE} = 4.795$); (C) self-sustained oscillations ($\omega_{EE} = 5.2$). Left-hand panels throughout present the resulting rotation numbers $\rho$. of the stable solutions. Right-hand panels show the average amplitude of the stable oscillator. White areas in panels (B) and (C) correspond to the solution existing outside of the defined domain. Baseline physiological parameters are stated in the Supplementary Materials. Harmonic response ($\rho = 1$, orange), and half-harmonic responses ($\rho = 1/2$, cyan) are the largest regions.

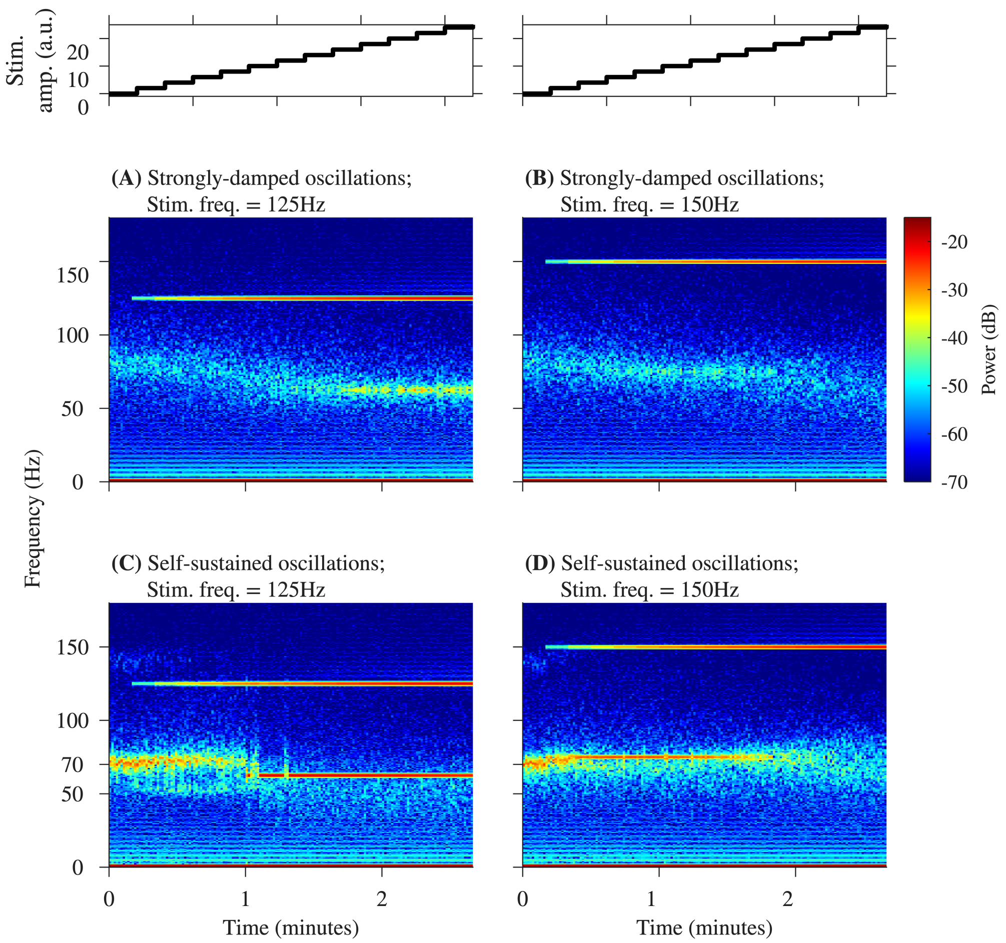


Figure S7: **Example predictions of a neuromodulation experiment with the inclusion of noise** Top panels show the time course of the stimulation amplitude as it is ramped up in discrete steps every 10 seconds. (A)-(D) spectrograms for four scenarios. Left panels (A, C): stimulation frequency 125 Hz. Right panels (B, D): stimulation frequency 150 Hz. Panels (A-B) show simulations of the system, with strongly-damped oscillations ($\omega_{EE} = 4.0$). (C-D) simulations in the system with self-sustained oscillations at 70 Hz, ($\omega_{EE} = 5.2$). Baseline physiological parameters are given in the Supplementary Materials.

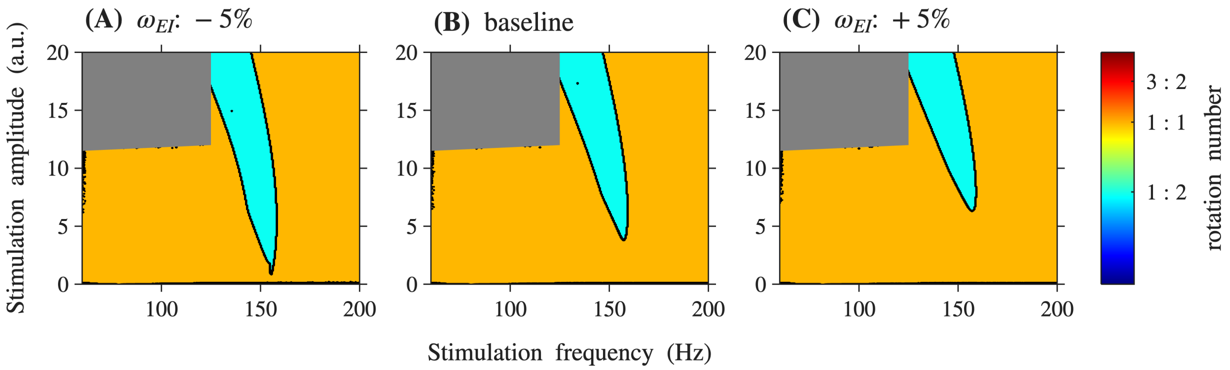


Figure S8: **Changes in tongue structure of weakly-damped system due to perturbations of parameter** $\omega_{EI}$ Oscillatory behaviour as a function of the stimulation amplitude and stimulation frequency. (A) Rotation numbers recorded when the baseline parameter value was decreased by 5%. (B) Rotation numbers recorded with respect to baseline parameter values. (C) Rotation numbers recorded when the baseline parameter value was increased by 5%.

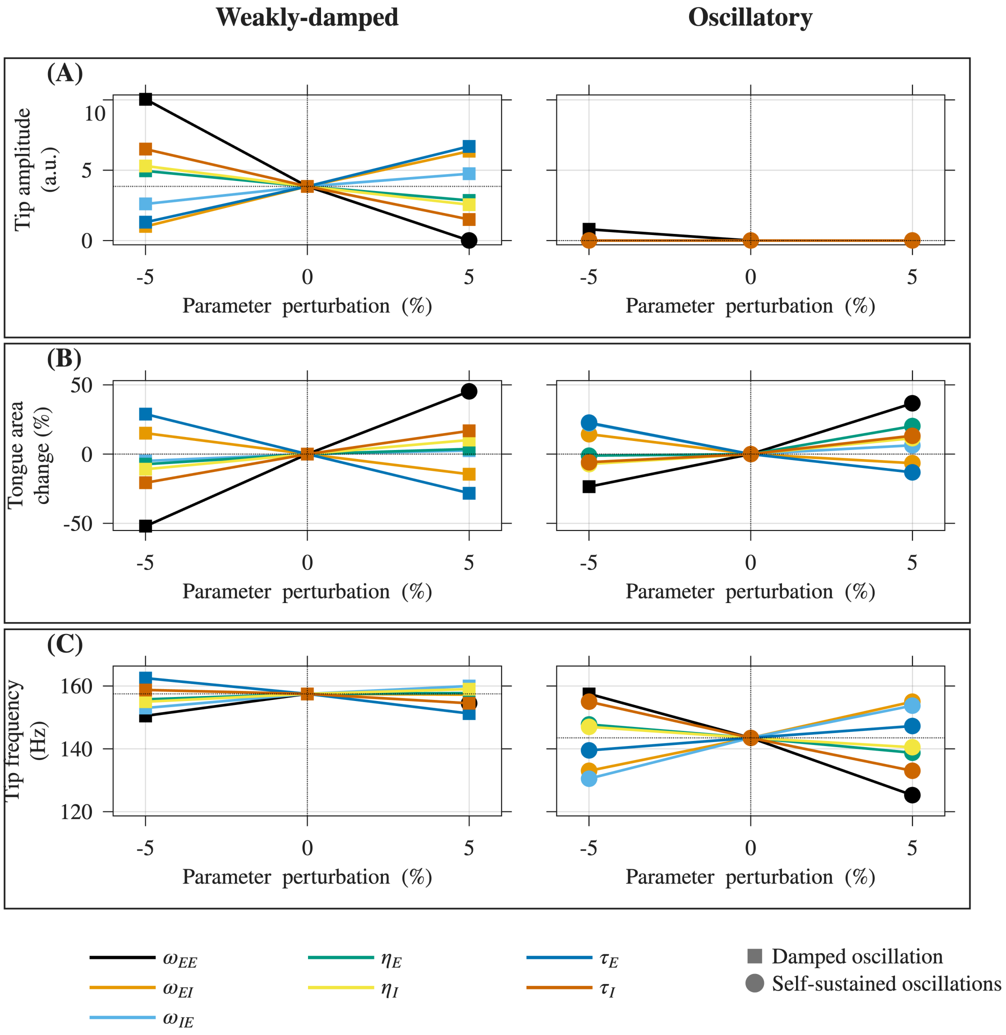


Figure S9: **Metrics of sensitivity for individual parameter changes in the case of weakly-damped and self-sustained oscillations** All metrics are taken with respect to the 1:2 tongue. In all panels, the 0% case demonstrates the baseline measurement. Left panels correspond to changes in the weakly-damped system, right panels correspond to the changes in the system with self-sustained oscillations. (A) Location of the tip of the 1:2 tongue in the amplitude dimension. (B) Change of the 1:2 tongue area compared to the baseline measurement. (C) Location of the tip of the 1:2 tongue in the frequency dimension.

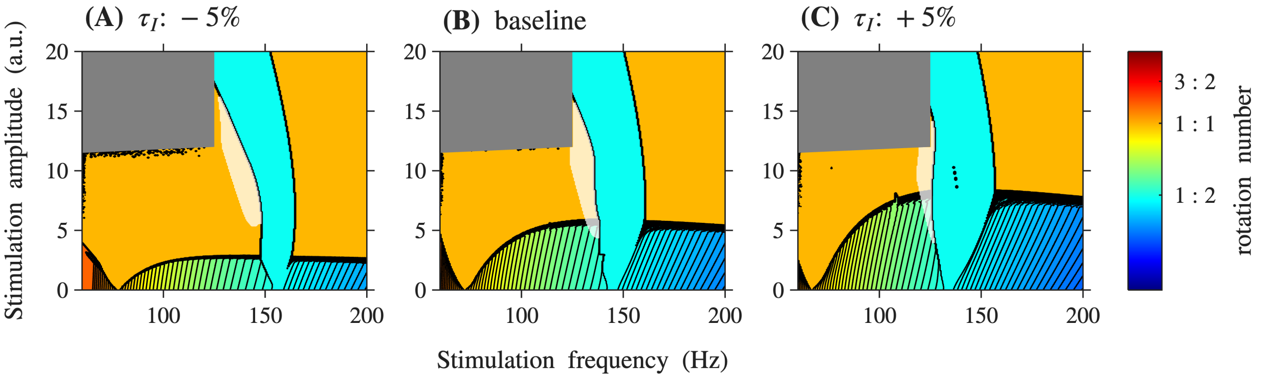


Figure S10: **Changes in tongue structure of the system with self-sustained oscillations due to perturbations of parameter** $\tau_I$ Oscillatory behaviour as a function of the stimulation amplitude and stimulation frequency. Hysteresis area is marked with a grey overlay. (A) Rotation numbers recorded when the baseline parameter value was decreased by 5%. (B) Rotation numbers recorded with respect to baseline parameter values. (C) Rotation numbers recorded when the baseline parameter value was increased by 5%.

Table S4: Summary of the local sensitivity analysis. Each parameter was perturbed independently by ±5% around the representative parameter sets. Response classes are abbreviated as SD (strongly damped), WD (weakly damped), and Osc (self-sustained oscillations). Tick marks indicate that a particular behaviour was observed; cross marks indicate lack of thereof.

| **Baseline dynamical regime** | **Outcome** | $\omega_{EE}$ | | $\omega_{EI}$ | | $\omega_{IE}$ | | $\eta_E$ | | $\eta_I$ | | $\tau_E$ | | $\tau_I$ | |
|---|---|---|---|---|---|---|---|---|---|---|---|---|---|---|---|
| | | −5% | +5% | −5% | +5% | −5% | +5% | −5% | +5% | −5% | +5% | −5% | +5% | −5% | +5% |
| Strongly damped | Resulting class | SD | SD | SD | SD | SD | SD | SD | SD | SD | SD | SD | SD | SD | SD |
| | 1:2 tongue | ✗ | ✗ | ✗ | ✗ | ✗ | ✗ | ✗ | ✗ | ✗ | ✗ | ✗ | ✗ | ✗ | ✗ |
| | Other subharmonics | ✗ | ✗ | ✗ | ✗ | ✗ | ✗ | ✗ | ✗ | ✗ | ✗ | ✗ | ✗ | ✗ | ✗ |
| | Hysteresis | ✗ | ✗ | ✗ | ✗ | ✗ | ✗ | ✗ | ✗ | ✗ | ✗ | ✗ | ✗ | ✗ | ✗ |
| Weakly damped | Resulting class | WD | **Osc** | WD | WD | WD | WD | WD | WD | WD | WD | WD | WD | WD | WD |
| | 1:2 tongue | ✓ | ✓ | ✓ | ✓ | ✓ | ✓ | ✓ | ✓ | ✓ | ✓ | ✓ | ✓ | ✓ | ✓ |
| | Other subharmonics | ✗ | ✓ | ✗ | ✗ | ✗ | ✗ | ✗ | ✗ | ✗ | ✗ | ✗ | ✗ | ✗ | ✗ |
| | Hysteresis | ✗ | ✓ | ✗ | ✗ | ✗ | ✗ | ✗ | ✗ | ✗ | ✗ | ✗ | ✗ | ✗ | ✗ |
| Self-sustained oscillations | Resulting class | **WD** | Osc | Osc | Osc | Osc | Osc | Osc | Osc | Osc | Osc | Osc | Osc | Osc | Osc |
| | 1:2 tongue | ✓ | ✓ | ✓ | ✓ | ✓ | ✓ | ✓ | ✓ | ✓ | ✓ | ✓ | ✓ | ✓ | ✓ |
| | Other subharmonics | ✗ | ✓ | ✓ | ✓ | ✓ | ✓ | ✓ | ✓ | ✓ | ✓ | ✓ | ✓ | ✓ | ✓ |
| | Hysteresis | ✗ | ✓ | ✓ | ✓ | ✓ | ✓ | ✓ | ✓ | ✓ | ✓ | ✓ | ✓ | ✓ | ✓ |

### 6 Preliminary example of entrainment with $\rho = 1/4$ in a human participant

Here, we present illustrative LFP recordings from a PD patient in an off-medication state with a bilaterally implanted DBS device. The stimulation is delivered to the STN. Stimulation was applied at 125 Hz with 60 $\mu$s pulse width, increasing and decreasing stimulation amplitude. LFP recordings were taken at 250 Hz. The protocol and methods, including data processing methods, followed those reported in [2].

In the LFP recordings, we notice the existence of an endogenous rhythm at approximately 45-50 Hz, which is present off-stimulation. At certain stimulation frequencies (when the amplitude is increased from 1 mA to 2 mA and decreased back to 1 mA again), a 1/4-subharmonic response is seen at 31.25 Hz. The half-harmonic seen at 62.5 Hz should be treated as a superharmonic of the previous result.

We do not believe the quarter-harmonic to be an artefact, since it is not present during the stimulation at 1 mA but is present throughout stimulation between 1 and 2 mA. In contrast, the response at 18 Hz appears to be clearly artefactual, persisting throughout the recording, regardless of the stimulation settings.

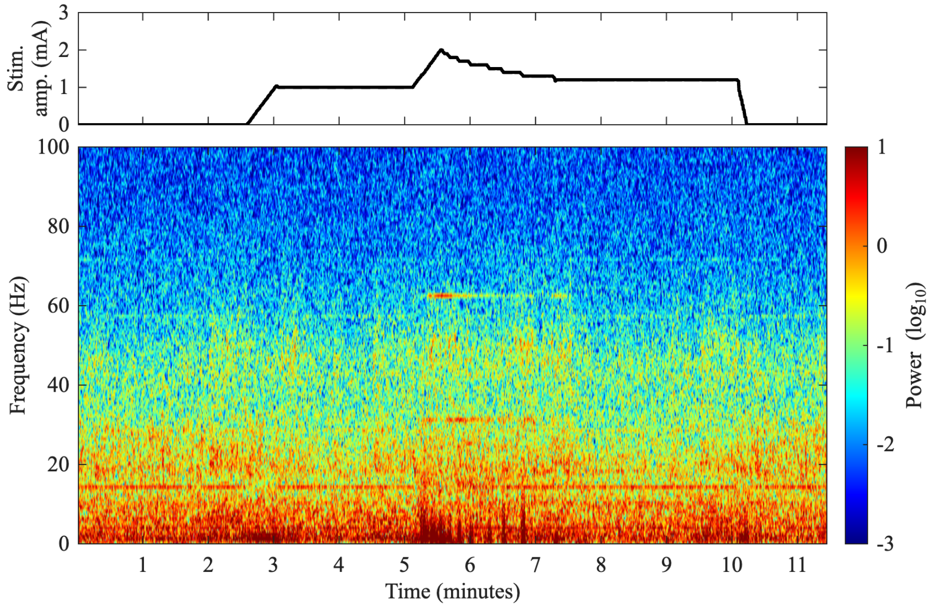


Figure S11: **Example of 1/4-subharmonic response recorded in a human participant.** Spectrogram of the LFP recording from a patient living with PD. In the clinical protocol, recording started off stimulation, with the amplitude gradually increased after that time. The top panel displays the changes in the amplitude. 1/4-subharmonic response is seen between 5-7 minutes of the recording. For details about the methodology and signal processing, see [2].

### 7 Non-autonomous oscillators and Arnold tongues

In the presence of stimulation, equations (S1) are non-autonomous. The phenomenon of entrainment refers to an interaction between two or more oscillators. Unlike in models of synchronisation, where oscillators affect each other's dynamics, entrainment models consider scenarios where one periodic oscillator acts as a forcing term whose characteristics remain unaffected by the system's overall dynamics. The forcing, characterised by its frequency (or period) and strength (such as amplitude), drives the system to become phase-locked with the external periodic input.

While most common, it is not necessary that the entrained oscillation has the same period as the one of the forcing rhythm (i.e., one oscillation of the system for each oscillation of the drive). The nature of the phase-locking is characterised by the *rotation number*.

Within the frequency-amplitude space, system responses of a particular rotation number form areas, referred to often as *Arnold Tongues* [1]. One of the intricacies of the tongue structure is that there exists a tongue corresponding to each rational rotation number. These tongues have their tips located at every rational number on the frequency axis – meaning that there are an infinite number of tongues. As the amplitude of the forcing increases, some tongues cease to exist, and the nature of the phase-locking changes, ultimately leading to dichotomies and chaotic solutions [5].

##### Entrainment of endogenous rhythms and the structure of the tongues

Throughout this work, we have presented the structure of the entrainment tongues with regard to the changing stimulation frequency. More generally, however, the structure of the tongues depends on the ratio of natural and forcing frequencies, or (equivalently) the ratio of the forcing and natural periods. We illustrate this in Figure S12. In particular, the tips of the entrainment tongues are located at specific values. The tip of the 1:1 tongue is located where the period of the forcing matches that of the oscillator. The tip of the 1:2 tongue is located where the period of the forcing is 1/2 of the natural oscillation period. In general terms, the tip of the tongue with rotation number $\rho = p/q$ is located where the period of the forcing is at the $p/q$ ratio with the oscillator's natural period.

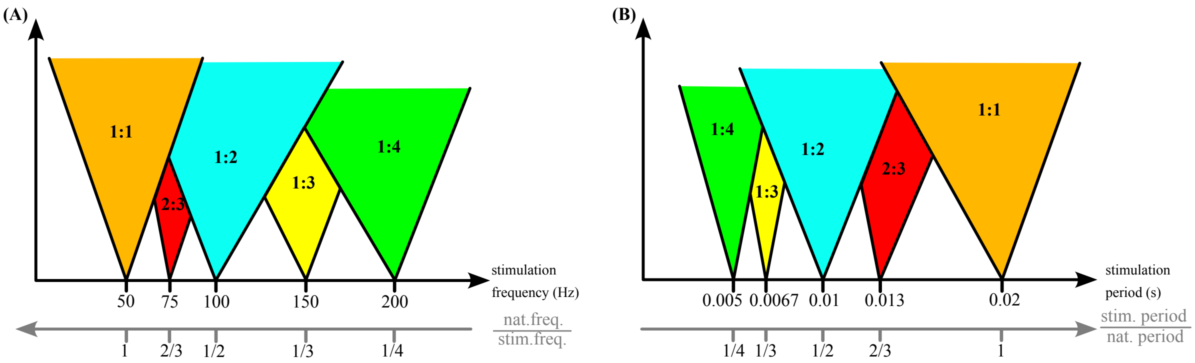


Figure S12: **General structure of Arnold Tongues**. Diagram for illustration purposes only, not to scale. The two pictures are equivalent, with the stimulation frequency changed in (A) and the stimulation period changed in (B). Note that the position of the tongue's tip is determined by the ratio of the forcing frequency (period) and the natural frequency (period) of the oscillator.
